# A genome-wide association analysis of 2,622,830 individuals reveals new pathogenic pathways in gout

**DOI:** 10.1101/2022.11.26.22281768

**Authors:** Tanya J. Major, Riku Takei, Hirotaka Matsuo, Megan P. Leask, Ruth K. Topless, Yuya Shirai, Zhiqiang Li, Aichang Ji, Murray J. Cadzow, Nicholas A. Sumpter, Marilyn E. Merriman, Amanda J. Phipps-Green, Mariana Urquiaga, Eric E. Kelley, Rachel D. King, Sara E. Lewis, Brooke A. Maxwell, Wen-Hua Wei, Sally P.A. McCormick, Richard J. Reynolds, Kenneth G. Saag, Matthew J. Bixley, Tayaza Fadason, Justin M. O’Sullivan, Lisa K. Stamp, Nicola Dalbeth, Abhishek Abhishek, Michael Doherty, Edward Roddy, Lennart T.H. Jacobsson, Meliha C. Kapetanovic, Olle Melander, Mariano Andrés, Fernando Pérez-Ruiz, Rosa J Torres, Timothy Radstake, Timothy L. Jansen, Matthijs Janssen, Leo A.B. Joosten, Ruiqi Liu, Orsi Gaal, Tania O. Crişan, Simona Rednic, Fina Kurreeman, Tom W.J. Huizinga, René Toes, Frédéric Lioté, Pascal Richette, Thomas Bardin, Hang Korng Ea, Tristan Pascart, Geraldine M. McCarthy, Laura Helbert, Blanka Stibůrková, Anne-K. Tausche, Till Uhlig, Véronique Vitart, Thibaud S. Boutin, Caroline Hayward, Philip L. Riches, Stuart H. Ralston, Archie Campbell, Thomas M. MacDonald, the FAST Study, Akiyoshi Nakayama, Tappei Takada, Masahiro Nakatochi, Seiko Shimizu, Yusuke Kawamura, Yu Toyoda, Hirofumi Nakaoka, Ken Yamamoto, Keitaro Matsuo, Nariyoshi Shinomiya, Kimiyoshi Ichida, Japan Gout Genomics Consortium, Chaeyoung Lee, Linda A. Bradbury, Matthew A. Brown, Philip C. Robinson, Russell R.C. Buchanan, Catherine L. Hill, Susan Lester, Malcolm D. Smith, Maureen Rischmueller, Hyon K. Choi, Eli A. Stahl, Jeff N. Miner, Daniel H. Solomon, Jing Cui, Kathleen M. Giacomini, Deanna J. Brackman, Eric M. Jorgenson, 23andMe Research Team, Wei Wang, Suyash Shringarpure, Alexander So, Yukinori Okada, Changgui Li, Yongyong Shi, Tony R. Merriman

## Abstract

Gout is a chronic disease of monosodium urate crystal deposition in the setting of hyperuricemia that typically presents with recurrent flares of acute inflammatory arthritis that occur due to innate immune response to deposited crystals. The molecular mechanism of the progression from hyperuricemia to clinical gout is poorly understood. Here we provide insights into this progression from a genetic study of 2.6 million people, including 120,282 people with gout. We detected 376 loci and 410 genetically independent signals (148 new loci in urate and gout). We identified 1,768 candidate genes with subsequent pathway analysis revealing urate metabolism, type 2 diabetes, and chromatin modification and structure as top pathways in gout. Genes located within or statistically linked to significant GWAS loci were prioitized for their potential to control the progression from hyperuricemia to gout. This identified strong candidate immune genes involved in epigenetic remodelling, cell osmolarity, and regulation of NLRP3-inflammasome activity. The genetic association signal at *XDH*, encoding the urate-producing enzyme xanthine oxidoreductase (XOR), co-localizes with genetic control of *XDH* expression, but only in the prostate. We demonstrate XOR activity and urate production in the mouse prostate, and use single-cell RNA sequence data to propose a model of urate reuptake, synthesis, and secretion by the prostate. The gout-associated loci were over-represented for genes implicated in clonal hematopoeiesis of indeterminate potential (CHIP) and Mendelian randomization analysis provided evidence for a causal role of CHIP in gout. In concert with implication of epigenomic regulators, this provides support for epigenomic remodelling as causal in gout. We provide new insights into the molecular pathogenesis of gout and identify an array of candidate genes for a role in the inflammatory process of gout.

## Introduction

Gout is an important public health issue of increasing prevalence^1^ and burden^2^ that is exacerbated by co-morbidity with cardiometabolic and renal disease^3^. Prevalence is approximately four-fold greater in men than in women^1^. It typically presents as recurrent self-limiting attacks of extremely painful acute inflammatory arthritis (termed ‘gout flares’). Gout is a chronic disease of monosodium urate (MSU) crystal deposition in joints and other structures that occurs in the setting of elevated serum urate levels (hyperuricemia)^4^. The gout flare is initiated when MSU crystals interact with resident macrophages to form and activate the NOD-like receptor protein 3 (NLRP3) inflammasome, which results in production of mature IL-1β and IL-18^5^. The inflammatory response is then amplified by the recruitment and activation of innate immune cells such as neutrophils, with the resolution phase mediated by processes including anti-inflammatory cytokines and aggregated neutrophil extracellular traps^6^. Soluble urate trains the immune system^7^ to have increased activity and response to MSU crystals through durable epigenetic modifications^8^.

Large genome-wide association studies (GWAS) have provided insight into the molecular mechanisms of urate control, with genetic variation in loci containing renal and intestinal urate transporters being prominent (e.g. *SLC2A9, ABCG2, SLC22A12*), with the kidney and liver the main organs in which genetic control is exerted^9^. In contrast, GWAS in gout have been relatively small^9–15^, the largest studies comprise 13,179^9^ and 37,105^16^ individuals with gout and identified 27 and 52 gout-associated genes, respectively; the majority of which also associate with urate levels. Whilst understanding the genetic and molecular control of serum urate is important in gout, most individuals with hyperuricemia do not develop gout (only up to 36%^17,18^) and many individuals with hyperuricemia who have not experienced a gout flare have imaging-confirmed monosodium urate crystal deposits.^19^ Subclinical inflammation is associated with these deposits, indicating other factors must be involved in developing symptomatic gout^20^. Estimates of serum urate heritability from twin studies range from 45 to 73%, but there are no credible estimates of the heritability of gout based on twin studies^20^. Heritability calculated using genome-wide genotype data in a cohort of unrelated European individuals was estimated at 28 to 30%, which is consistent with a recently-reported heritability estimate (27.9%) of clinically-defined gout in Japanese populations^21^, suggesting gout may be less heritable than hyperuricemia. Nevertheless genetic studies should be able to elucidate mechanisms involved in progressing from hyperuricemia to gout^22^.

There is some evidence for genetic variants controlling the progression from hyperuricemia to gout. GWAS in gout comparing to individuals with asymptomatic hyperuricemia have identified a small number of loci^10,23,24^ (most of which also associate with serum urate levels^9,25^), and a Polynesian-specific variant in *IL-37* associates with gout compared to asymptomatic hyperuricemia^26^. However, these loci cannot necessarily be interpreted as controlling the progression from hyperuricemia to symptomatic gout because hyperuricemia is defined by a single urate measure in each study, which does not allow accounting for the lifetime burden of urate^27^. Candidate gene studies have associated variants in inflammatory genes with gout, but none are widely replicated^20^. We present findings from the largest GWAS in gout to date utilizing 120,282 cases and 2,502,548 controls over four ancestral groups. The findings illuminate previously unidentified candidate pathways with the potential to control the progression from hyperuricemia to clinical gout.

## Results

### Study overview

The study design is summarized in **Figure S1**. This study comprised GWAS in four ancestral groups (African, East Asian, European, Latinx) and a trans-ancestry meta-analysis, each including sex-specific analyses. Summary statistics were amalgamated from 120,282 people with and 2,502,548 people without gout. The effect of a polygenic risk score on risk and prediction of gout was evaluated, as was genome-wide genetic correlation with other phenotypes. Follow-up analyses included identification of candidate genes by co-localizing signals of association with expression quantitative trait loci (eQTL), identifying genes with candidate missense variants, and genes identified by MAGMA^28^. These candidate genes were applied to pathway analysis. A prioritization scheme identified candidate genes for the progression from hyperuricemia to gout. Mechanistic studies included investigation of the role of xanthine oxidoreductase in the prostate in synthesizing urate, and the evaluation of a causal role for the clonal hematopoiesis of indeterminate potential (CHIP) pathway in gout.

### Identification of GWAS signals

#### Single- and trans-ancestry genome-wide analyses

We carried out fixed-effect inverse-variance weighted GWAS meta-analyses for gout in four ancestral groups (African (AFR) – 3,052 cases and 77,891 controls; East Asian (EAS) – 10,729 cases and 82,807 controls; European (EUR) – 100,661 cases and 2,106,003 controls; and Latinx (LAT) – 5,840 cases and 235,847 controls) (**Table S1**). The four GWAS were conducted using imputed genotypes that included only biallelic single nucleotide polymorphisms (SNPs) with a minor allele frequency > 0.1% (**Methods** and **Table S1**). The African meta-analysis included 24,649,233 SNPs, the East Asian meta-analysis included 10,194,540 SNPs, the European meta-analysis included 13,443,610 SNPs, and the Latinx meta-analysis included 18,956,225 SNPs. A trans-ancestry meta-analysis of the variants present in all four ancestry-specific GWAS (6,386,511 SNPs) was also conducted to maximize locus discovery by increased sample size, and it comprised 120,282 cases and 2,502,548 controls. Whilst there was some indication of inflation of GWAS test statistics (IJ_GC_ = 1.050_AFR_, 1.121_EAS_, 1.611_EUR_, 1.105_LAT_), linkage disequilibrium (LD) score regression analysis revealed no evidence of inflation due to factors such as population structure (LD score intercept = 1.025_AFR_, 1.049_EAS_, 1.026_EUR_, 1.048_LAT_), with the elevated IJ_GC_ observed in the European GWAS likely being due to polygenicity between individual cohorts contributing to the ancestry-specific meta-analysis^29^.

#### Identification of significant loci and independent signals

A total of 296 genome-wide significant loci (defined as a genomic segment with ≥1 lead variant(s) that define a genetically independent signal) were detected in the trans-ancestry meta-analysis (log_10_BF ≥ 6), 20 with >1 genetically-independent signal, resulting in 318 independent genome-wide significant signals. In the separate ancestry GWAS, two genome-wide significant loci were detected in the African GWAS, 10 in the East Asian GWAS, 277 in the European GWAS (13 with multiple signals resulting in 291 independent lead SNPs), and ten in the Latinx GWAS (**Figure 1; Figure S1, S2; Table S2**). There was a total of 339 non-overlapping loci and 365 genetically independent signals across all analyses. Only two loci were common to all ancestral-specific analyses (**Figure 1**), both on chromosome 4 and containing the well-studied *SLC2A9* and *ABCG2* genes^20^. The *ABCG2* locus had the strongest statistical evidence for association with gout in the East Asian, European, Latinx, and trans-ancestry GWAS. A total of 119 loci were unique to one of the five GWAS conducted. Where a locus was identified across more than one of the five GWAS, but there was a different lead SNP between the ancestries, the majority of the divergent lead SNPs were in high linkage disequilibrium (r^2^ ≥ 0.8; 93 of 139 loci; **Table S2**). For the identified lead variants, there was strong correlation between effects on serum urate level in the UK Biobank and gout risk (Pearson correlation *r* = 0.92; **Figure S3**).

**Figure 1:**
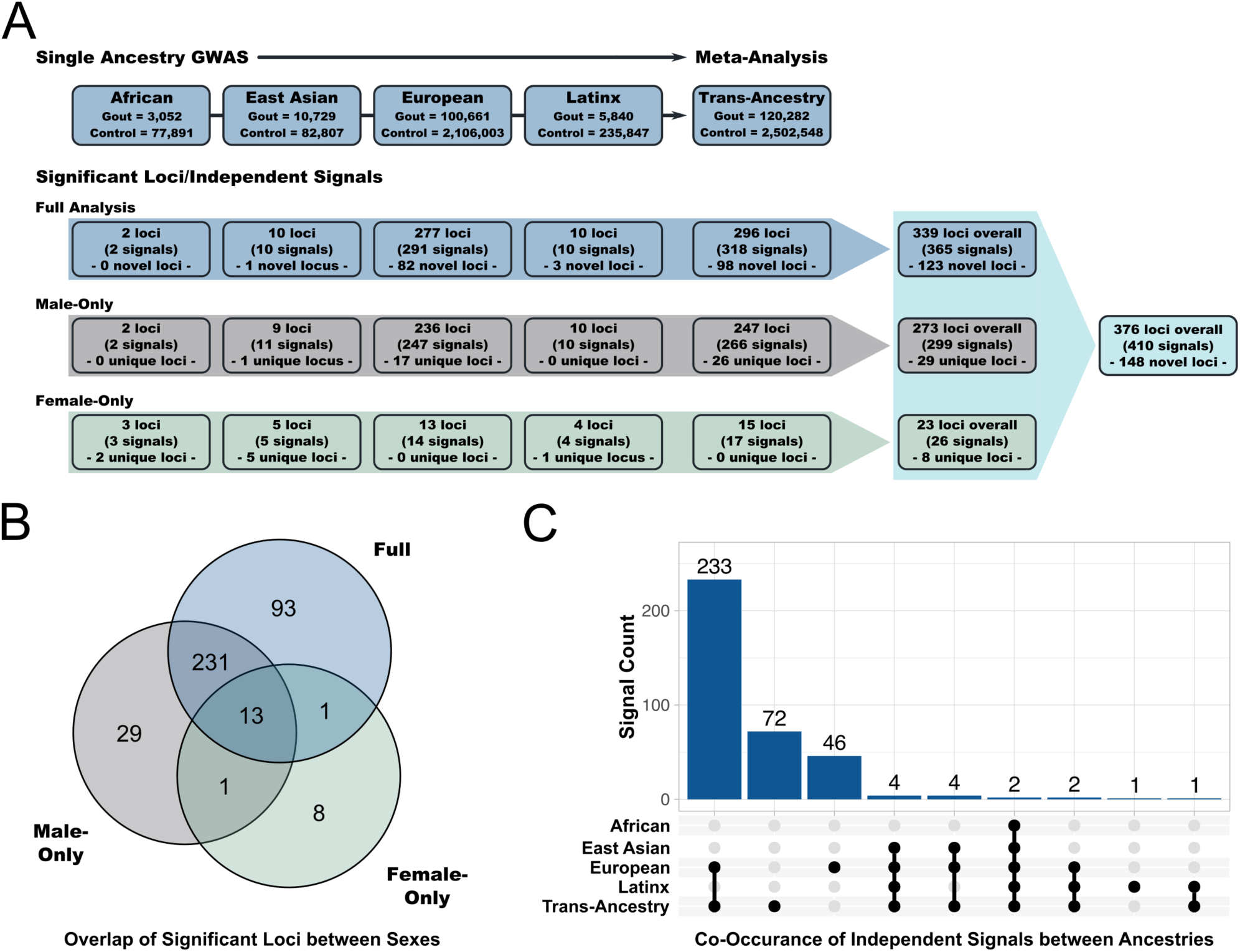
Numbers of significant loci and independent signals, across the ancestry-specific and trans-ancestry analyses and in the full (combined sexes), male-only, and female-only analyses. **A)** summarizes the cohorts used, the GWAS done, and loci detected. **B)** shows a Venn diagram of the overlap between significant loci across the full, male-only, and female-only GWAS amalgamated across all ancestries. **C)** shows an upset plot of the overlap between the significant independent signals in the single-ancestry and trans-ancestry analyses for the full cohort.

#### Sex-stratified analyses

The prevalence of gout is at least three to four-fold greater in men than women^1,30^. Therefore, we conducted sex-stratified GWAS for each of the four ancestral groups and the trans-ancestry meta-analysis. In each ancestry-specific analysis there were fewer female gout cases than male gout cases (ranging from a 1:2 ratio to a 1:23 ratio of women to men; **Table S1**). A total of 247 loci were identified in the male-only trans-ancestry meta-analysis (19 with multiple signals resulting in 266 independent genetic associations) and 15 significant loci were identified in the female-only trans-ancestry meta-analysis (17 independent signals) (**Table S2**). Across all ancestries there was a total of 299 independent signals in the male-only analyses and 26 independent signals in the female-only analyses; 29 of the male-only loci (36 signals) and 8 of the female-only loci were not detected in the full (combined sexes) analyses. One locus was detected in both the male-only and female-only analyses, but not the full (combined sexes) analysis (**Figure 1**). This was due to heterogeneity between the male-only and female-only signals (**Table S4**).

Of the 410 genetically independent signals identified across all analyses, 44 (representing 36 loci) had significant heterogeneity (*P* < 1×10^−6^) between the male-only and female-only results, with effect sizes greater in men for all 36 loci (**Table S3**). Thirty-two (89%) had previously been associated with urate^9^ and include the 10 loci with the strongest effects on urate^31^ (*SLC2A9, ABCG2, SLC17A1, SLC16A9, GCKR, SLC22A11, INHBC, RREB1, PDZK1, SLC22A12*). *ABCG2* was heterogenous in all datasets excepting African, and *SLC2A9* was heterogenous in the trans-ancestry meta-analysis and European GWAS. Of the 32 loci previously associated with urate, 13 had also been reported as exhibiting heterogeneity between sexes in urate control^9^, with ten having a stronger effect in men and three in women (*SLC2A9, A1CF, IGF1R*).

#### Previously unreported loci

Of the 376 loci identified across all analyses, 148 (four with two independent signals) have not previously had any variant within the locus boundaries associated with urate or gout. Thus, these 148 loci can be considered newly identified associations with gout (**Table S4**). Of the remaining 228 loci, 143 had the same lead SNP or were in high LD (r^2^ ≥ 0.8), 64 in low to moderate LD (0.1 ≤ r^2^ < 0.8), and 21 were in no linkage disequilibrium (r^2^ < 0.1) with a SNP previously reported to be associated with serum urate or gout (**Table S4**). Of the 30 loci with >1 independent signal, 18 loci (19 independent signals) had no linkage disequilibrium (r^2^ < 0.1) between the previously reported SNP(s) at the locus and one of the independent lead SNPs. Thus, 188 (148 + 21 + 19) of the independent signals can also be considered newly identified gout associations.

### Risk prediction and genetic relationships

#### Polygenic risk score and risk of gout

Gout polygenic risk scores (PRS) consisting of 289 SNPs from the European and 316 from the trans-ancestry GWAS, weighted by effect size, were generated and applied to the UK Biobank European-ancestry cohort to assess how well the GWAS results predict gout status (**Table S5**). The risk score values were split into ten bins with equal ranges, and gout prevalence increased across these ten risk score bins from 0.0 to 24.3% and 0.0 to 41.7%, respectively for the European and trans-ancestry scores **(Figure 2; Figure S4; Table S5**). Compared to the risk score bin that included the most individuals, age- and sex-adjusted odds-ratios (ORs) ranged from 0.22 [95% CI 0.11; 0.45] and 0.07 [0.02; 0.28], for European and trans-ancestry respectively, in the lowest risk score bin, to 20.7 [9.2; 46.8] and 50.0 [20.4; 122.6], respectively, in the highest risk score bin. For comparison, male sex conferred a gout risk of 13.9 [12.7; 15.1]. Male prevalence of gout in the highest European and trans-ancestry risk score bins was 39.1% (OR = 23.7 [10.1; 55.4]) and 58.8% (OR = 60.7 [22.7; 162.5]), respectively. While there were no females with gout in the highest bin for either risk score, analysis of the highest bin with gout cases gave OR = 9.4 [3.8; 23.3] and OR = 6.2 [2.3; 17.0] for European and trans-ancestry risk scores, respectively.

**Figure 2:**
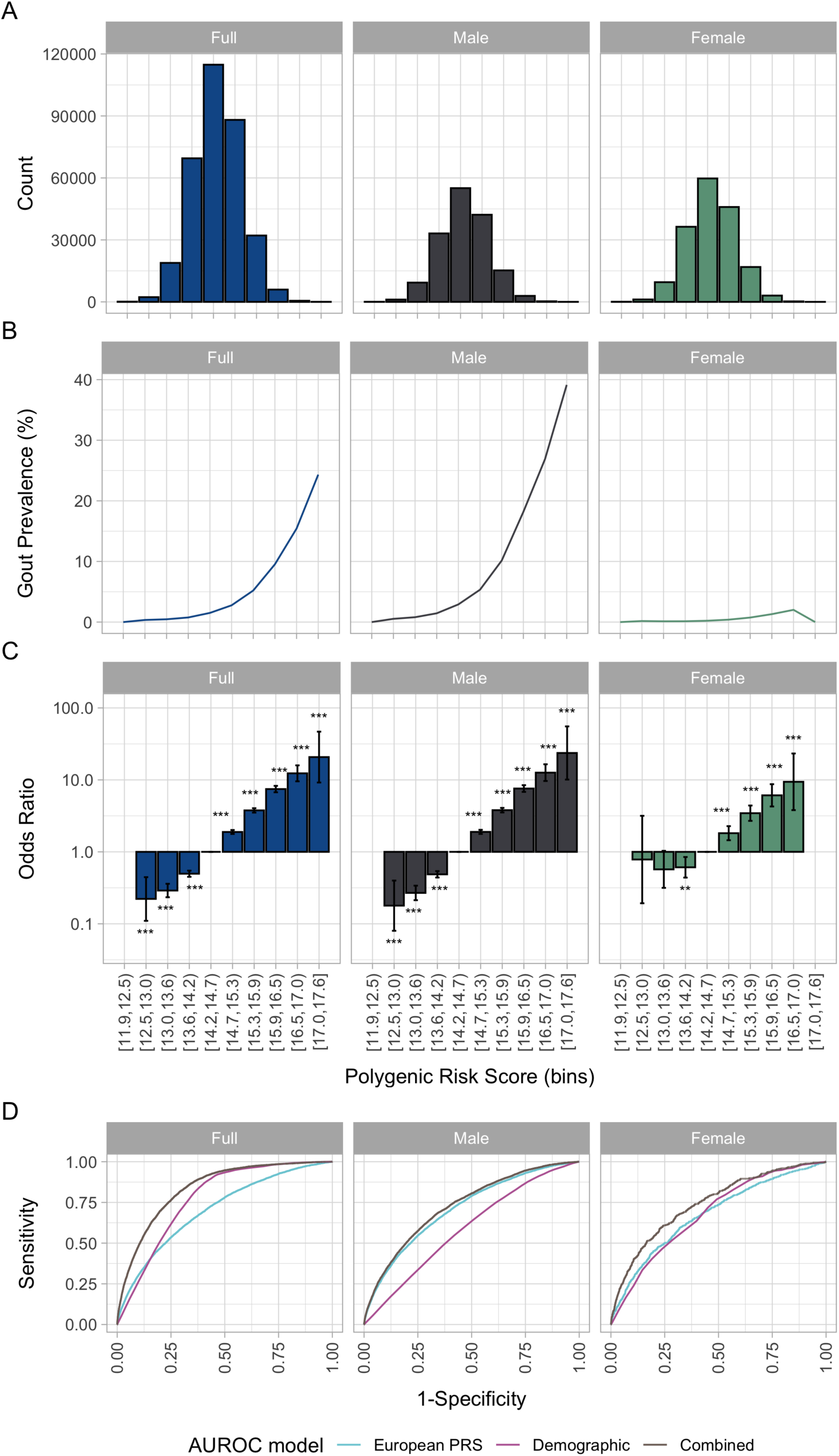
Association of polygenic risk score with gout in European participants of the UK Biobank in combined sexes, men, and women. **A)** shows the polygenic risk score bin distribution, **B)** shows the gout prevalence in different polygenic risk score bins, **C)** shows the risk of gout for each different risk score bin, compared to the most common bin (as visualized in panel A), and **D)** shows the area under the receiver operating characteristic curve graphs.

Equivalent risk scores were generated using SNPs from the European and trans-ancestry male-only and female-only GWAS. The 246-SNP risk score derived from the male-only European GWAS had a higher prevalence of gout (50.0%) and higher risk of gout (OR = 36.4 [2.2; 609.3]) in the highest risk score bin than the highest risk score bin generated from the full (sexes combined) European GWAS SNPs (**Figure S5**). The 265-SNP risk score derived from the male-only trans-ancestry GWAS did not improve upon the full trans-ancestry risk score in the highest risk score bin (gout prevalence 26.9%; OR = 15.1 [6.2; 36.3]). Both the European and trans-ancestry female-only risk scores had higher gout prevalence (5.3% and 3.8%) and OR (12.6 [3.0; 53.6] and 12.4 [3.8; 40.1]), respectively, in the highest bin than the risk score derived from the respective full (sexes combined) GWAS (**Figure S4**).

The 289-SNP European gout risk score improved gout risk prediction in Europeans (area under the receiver operating characteristic curve (AUROC) estimate of 0.71), compared to a 114-SNP risk score generated from a serum urate GWAS in European individuals^9^ (AUROC estimate of 0.66) (**Figure S6; Table S5**). These estimates were the same in the male-only AUROC estimates, and lower in the female-only estimates (0.68 for the gout risk score and 0.62 for the urate-derived risk score). Adding age increased the prediction accuracy of the gout risk score to 0.73 in males and 0.74 in females.

#### Genetic relationships with other phenotypes

To investigate the genetic correlation between gout and other complex phenotypes, LD score regression was undertaken using 934 traits reported in the UK Biobank, the European gout GWAS data and, for comparison, data generated from a GWAS of serum urate in European UK Biobank participants without gout (**Methods**). There was significant genetic correlation between gout and 348 phenotypes, which spanned 25 out of 27 broad phenotype categories (**Figure 3, Figure S7, Table S6**). As expected, the strongest correlations were with gout and urate (*r*_g_ ≥ 0.89). Outside of cardiovascular disease and its body composition-related risk factors, established as associated with gout, negative correlations were seen with two sex-related blood biomarkers, testosterone (*r*_g_ = −0.17) and sex hormone-binding globulin (*r*_g_ = −0.31). This is consistent with a recent report of greater genetically predicted levels of each hormone associating with reduced risk of gout^32^. There was positive genetic correlation between gout and blood counts of leukocytes, lymphocytes, neutrophils, eosinophils, and reticulocytes (*r*_g_ = 0.17, 0.14, 0.15, 0.12, and 0.26, respectively). Finally, positive genetic correlations between gout and various measures of negative mood were observed. The genetic correlations with urate levels were very similar (**Figure 3**) with 300 of the 348 phenotypes correlated with gout also significantly correlated with urate in the UK Biobank in a consistent direction.

**Figure 3:**
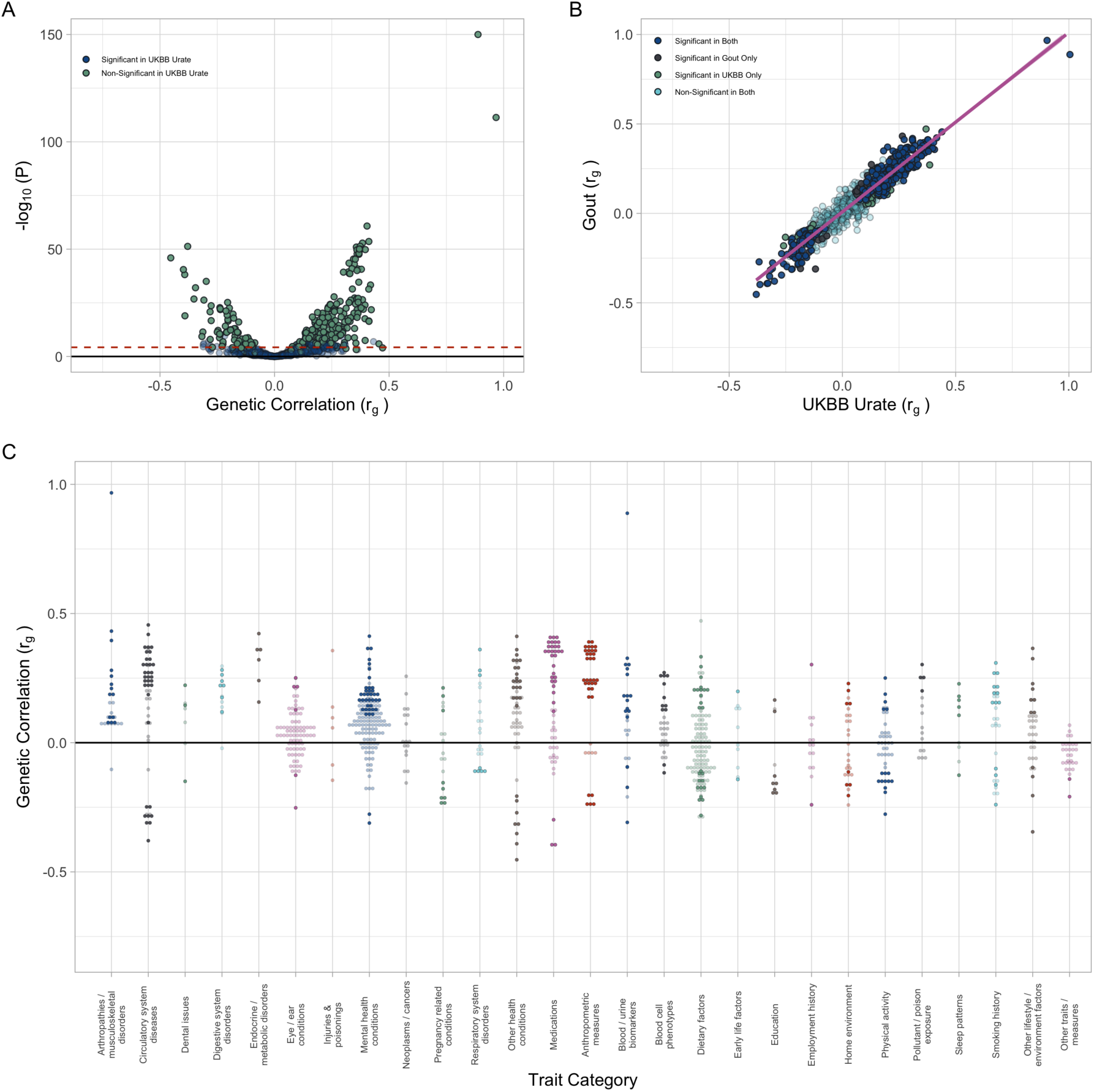
Genetic correlation between the European gout GWAS and UK Biobank GWAS traits. **A)** Volcano plot of genetic correlation between our European gout GWAS results and 934 UK Biobank GWAS traits. Points are coloured based on whether the equivalent correlation between our UK Biobank serum urate GWAS and the trait of interest was significant (*P* < 5.35×10^−5^). **B)** linear relationship between the genetic correlation results of our European gout GWAS and our UK Biobank serum urate GWAS. Points are coloured based on whether they were significant in both, one, or neither of the genetic correlation analyses. **C)** genetic correlation values (*r*_g_) across 27 trait categories. In all plots, each point represents the result of a genetic correlation analysis between our European gout GWAS and one of the 934 UK Biobank GWAS traits. Non-significant results are shown as transparent plot points for clarity.

To investigate the possibility of testing for causality of gout for the 348 phenotypes correlated with gout, we tested the 289 SNPs that were included in the European gout polygenic risk score (above) for evidence of vertical pleiotropy with the 348 phenotypes using two-sample Mendelian randomization (MR) in MRBase by the MR-Egger (Egger intercept) approach. There was extensive pleiotropy, with 69 phenotypes having evidence of pleiotropy at a Bonferroni-corrected level of significance (0.05/348 = 1.4×10^−4^), 93 with *P* < 0.001, and 146 with *P* < 0.01, which would complicate interpretation of any causal relationships identified via Mendelian randomization (**Table S7**). In the absence of a clear understanding of the molecular mechanisms of the genetic variants comprising the genetic instrument, further Mendelian randomization analysis was not pursued.

### Candidate Variants, Genes, and Tissues

#### Tissues enriched in gout

We analyzed the summary statistics of the four ancestries (African, East Asian, European, and Latinx) for significantly enriched tissues / cell-type groups and specific cell-types using covariate-adjusted LD score regression (cov-LDSC; Methods)^33^. In East Asian there was evidence of significant enrichment of tissues (Bonferroni-corrected *P*_coef_ ≤ 0.05/40) in kidney (10.8-fold enrichment; *P*_coef_ = 2.3×10^−4^), and in European there was enrichment in the kidney and liver (11.6-fold enrichment; *P*_coef_ = 8.6×10^−7^ and 5.1-fold enrichment; *P*_coef_ = 8.3×10^−6^, respectively) (**Table S8**; **Figure S8**). There were no significant enrichments observed in either of the African and Latinx analyses. With respect to which specific cell-types were significant in European, we observed significant cell-type associations (false discovery rate (FDR) ≤ 0.05) in small and large intestine, colonic mucosa, duodenum mucosa, rectal mucosa, pancreas, skeletal muscle, kidney, and liver (**Table S9; Figure S9**). There were three significant kidney cell histone marks found in the East Asian cell-type analysis (H3K4me3, H3K9ac, and H3K27ac) that were also identified in the European ancestry analysis (**Table S9**).

#### Cis- and trans-eQTL co-localization analyses

To identify candidate causal genes that are possibly controlled through regulation of gene expression, we focused on gout association signals that co-localized with *cis-* and *trans-*eQTL across 49 tissues in the Genotype Tissue Expression (GTEx) database. Of the 342 trans-ancestry and European lead SNPs present in GTEx (or its proxy if the lead variant was not present), 290 (84.8%) had at least one co-localized eQTL (posterior probability of co-localization (PPC) ≥ 0.5) representing 890 genes. There were 2,717 *cis*-eQTL over multiple tissues for 626 genes and 332 *trans*-eQTL (31 were both *cis*- and *trans*-eQTLs) over multiple tissues for 326 genes with evidence of co-localization (**Table S10; Figure S10**).

Of the 921 eQTL implicated by co-localization analysis, ten had control of expression by both a *cis-* and *trans*-eQTL (*ADK, HSP90AA1, RABEP2, SEPT9, SPN1, VPS9D1, RNF157, SLC16A9, TANC1* and *TUBB8P7*). For example, *rs1171614* is the lead SNP for a co-localized *cis*-eQTL for *SLC16A9* (encodes kidney monocarboxylate transporter SLC16A9 involved in carnitine transport) and *SLC16A9* is a *trans*-eQTL of *rs73592376* at the *CUBN* (proximal tubule uptake receptor) locus. We also noted that 47 of the 343 eQTL that had an eQTL in multiple tissues had opposing effects on gene expression between tissues (**Table S11**). Focusing on genes that may have a role in gout inflammation there were 11 genes with eQTLs in whole blood, of these genes five (*AQP10, BAG4, FADS1, SCAP, RP11-936I5.1*) had a direction of effect opposite to all other tissues in which an eQTL was observed.

Only five co-localized kidney cortex eQTL were observed in GTEx (*FGF5, HLF, ITIH4, RP11-307C18.1, NUDT2*), therefore we also leveraged comprehensive kidney eQTL data from the Susztaklab Kidney Biobank^34^ to identify additional kidney eQTL. We used LD (r^2^ > 0.8) between the lead gout and the lead kidney eQTL SNPs to identify 137 genes with gout-associated kidney eQTL, 69 of which also had a co-localized eQTL identified in other tissues within the GTEx data (**Table S12**).

We used Activity-By-Contact (ABC) data^35^ to determine if the lead signals with co-localized eQTL showed evidence of gene regulation through enhancers. Of the 290 lead variants that had signals co-localized with *cis*- and/or *trans*-eQTL, 55 variants were within an ABC enhancer for 386 genes with ABC-score ≥ 0.015 (**Table S13**). 31 of these 55 variants were within an activity-by-contact enhancer for 48 co-localized eQTL (all in *cis*; **Table S14**), indicating that these variants may be altering gene expression by directly affecting the functional interaction between the putative enhancer and gene promoter.

#### Tissue-specific eQTLs

Reasoning that co-localized eQTL restricted to a single GTEx-defined tissue may be more tractable to specific biological insights than those expressed in multiple tissues, we filtered for tissue-specific co-localized eQTL (**Table S15**), identifying 581 eQTL genes. Of specific interest are the eQTL in the testis for *SLC34A1, HNF4G,* and *PRPS2* – genes with known roles in urate transport, production, and homeostasis – suggesting that the function of these genes in urate control could have male-specific mechanisms. Additionally, the co-localized eQTL for *XDH* (encoding xanthine oxidoreductase (XOR)) was specific to the prostate. In female-specific tissues (ovary, vagina and uterus), we identified *COG5, JAZF1-AS1, FDX1P1, SPDYE18,* and *SDK1*, none of which play known roles in urate homeostasis or gout risk. We identified only two co-localized eQTLs specific to kidney tissue in GTEx (*FGF5* and *ITIH4*), and an additional 68 kidney eQTL from the Susztaklab Kidney Biobank^34^ not present in GTEx. Notable amongst the 68 was *SLC22A12* (encodes canonical urate transporter URAT1^36^) that has not previously been demonstrated to have gout-associated genetic control of expression in the kidney. We found seven liver-specific (*BSN, KLB, STK19B, TNKS, RAI1, ZDHHC14,* and *ZNF320*) and 16 whole blood-specific (*DGAT2, RPS6KB1, MBD5, C5orf42, MAP2K1, CDKL5, ARID4B, CARS, CSNK1G1, AUH, HEATR5B*, *AF064858.8, IL27, HAUS1P1, ZNF675* and *DPEP3*) co-localized eQTL.

#### DNA Methylation QTL co-localization analysis

The epigenetic reprogramming (‘trained immunity’) of monocytes by elevated levels of soluble urate is implicated in the etiology of gout^7,37,38^. To evaluate the possibility that gout-associated loci also associated with control of DNA methylation status in the blood, we conducted co-localization analysis between whole blood DNA methylation QTL (meQTL) in the GoDMC data set^39^ derived from European participants, using the 291 lead SNPs in the European GWAS. We identified 520 methylation CpG sites within 1Mb of a lead gout-associated SNP that also co-localized with a European GWAS signal (**Table S16**; PPC ≥ 0.8 and ≥ 100 variants in the 1Mb segment used for co-localization). To determine whether the implicated 520 CpG sites were involved in transcriptional regulation, we tested for enrichment in protein-binding within ±50 base pairs of the CpG sites using merged ChIP-seq data for 338 DNA-binding proteins from 1,536 cell-lines^40^. Significant enrichment was observed for 27 DNA-binding proteins (Bonferroni-corrected Fisher’s exact *P* ≤ 1.48×10^−4^) (**Table S17**) that bind to a total of 499 (96.0%) co-localized meQTL CpG sites (median = 54 CpG sites, mean = 76.9 CpG sites). Of the 27 DNA-binding proteins identified, only one was within a gout genetic association signal (*IRF1*). Amongst the top enriched DNA-binding proteins were the histone acetylation writers and readers EP300 and BRD4. BRD4 has previously been implicated in the regulation of the NLRP3 inflammasome through regulation of NFκΒ signalling,^41^ BRD4 inhibition ameliorates urate crystal-induced gouty arthritis by regulation of pyroptosis in a rat model^42,43^.

The kidney is a key organ in urate homeostasis. Given that variation in DNA methylation in the kidney mediates 46% of heritability in kidney function traits^44^, we also identified 328 genetic variants that were kidney meQTL for 576 CpG sites in strong LD (r^2^ ≥ 0.8) with a gout genetic association using the Liu *et al.*^44^ dataset (**Table S18**). The 50 kidney eQTL that also had at least one kidney meQTL were enriched for transcription factor binding of HNF4G using the enrichr gene set search engine^45^ (*P* = 0.03). For each of the kidney eQTL with co-localized meQTL we looked for SNPs in linkage disequilibrium (r^2^ ≥ 0.8) with the lead gout SNPs that overlap a transcription factor binding site for HNF4G using HaploReg. We found three variants that overlap and disrupt a matching HNF4A/G motif: *rs2453583* at *SLC47A1, rs1165183* at *H2BC1* (at the *SLC17A1-4* locus) and *rs260512* at *SKI*. Regulatory control of *SKI* and *H2BC1* has not previously been associated with gout or serum urate.

#### Fine-mapping of loci

We used three approaches (Bayes factor, FINEMAP^46^, and PAINTOR^47^) for fine-mapping the 376 unique loci in the European and trans-ancestry datasets from all analyses (full, male-only, and female-only. We considered 99% credible sets from any one of the separate approaches with at least one variant with posterior inclusion probability (PIP) ≥ 0.5. Between the three fine-mapping approaches and cohorts, we found a credible set with at least one variant with PIP ≥ 0.5 in 285 loci (76.6%) (**Table S19**). The fine-mapping of complex phenotype loci is compromised when summary statistics derived from meta-analysis of cohorts using different genotyping arrays and/or imputation panels are used. Therefore, we first identified and excluded compromised loci using SLALOM^48^. Of the 285 loci, 70 loci (24.6%) in total were flagged as compromised by SLALOM in at least one of the cohorts (**Table S20**). 215 loci remained after removing the compromised loci from the relevant cohort (**Table S21**) and 608 candidate causal SNPs (PIP ≥ 0.5 in one or more of the fine-mapping approaches and within a 99% credible set with ≤5 SNPs) were selected for further analysis (**Table S22**). Of these 608 variants, 37 were identified by two fine-mapping approaches and 16 were identified by all three approaches. We created a pool of 1,466 unique candidate causal variants (**Table S24**) from; a) the 799 lead SNPs from single- and trans-ancestry GWAS in all cohorts (254/799 = 31.8% present in the fine-mapping credible set results); b) from an additional 185 variants identified as conditionally associated with gout after applying GCTA-COJO^49^ to the European GWAS data (Methods; **Table S23**) (26/185 = 14.1% present in the fine-mapping credible sets); and c) the 608 variants identified by fine-mapping (**Table S22**) (there were 126 variants common to the three categories). The 1,466 SNPs in this pooled list were then assessed for their potential to disrupt gene function.

#### Candidate causal missense variants

Of the 1,466 SNPs in the pooled list, there were 21 missense variants. There were an additional 173 missense variants that were in strong LD with the 1,466 candidate variants (r^2^ > 0.8 in any of the relevant ancestries from 1000 Genomes) (**Table S25**). In addition to the genes with the 21 missense variants there were 26 other genes with a missense variant that was a lead SNP in one of the GWAS (including lead SNPs identified by conditional analysis) or had LD of r^2^ ≥ 0.98 with a lead SNP (**Table 1**). Of the combined 47 genes there were 16 with missense variants with information on protein function impact that supports causality, including 7 not previously reported as genetically or functionally implicated in serum urate control or gout (*ABCA6* p.Cys1359Arg, *GLS2* p.Leu581Pro, *MC4R* p.Val103Ile, *PNPLA3* p.Ile148Met, *SH2B3* p.Trp262Arg, *SLC39A8* p.Ala391Thr, *SLCO1B1* p.Val174Ala). Two of the genes with missense variants associated with serum levels of amino acids (*CPS1* and *GLS2*), and the *CUBN* and *LRP2* genes (encoding cubilin and megalin, respectively) physically interact in the kidney proximal tubule to resorb proteins from filtered urine^50^.

**Table 1:**
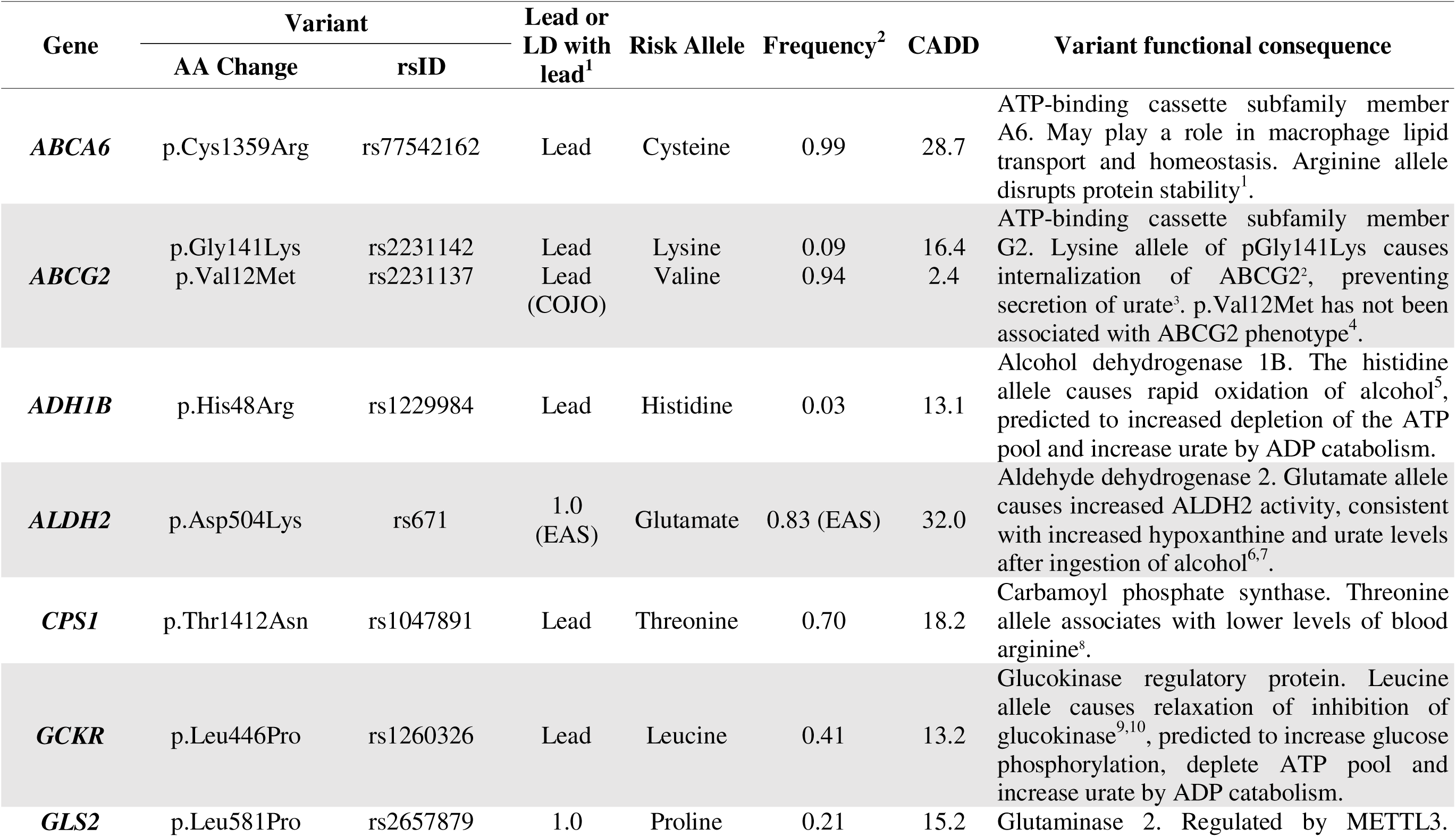
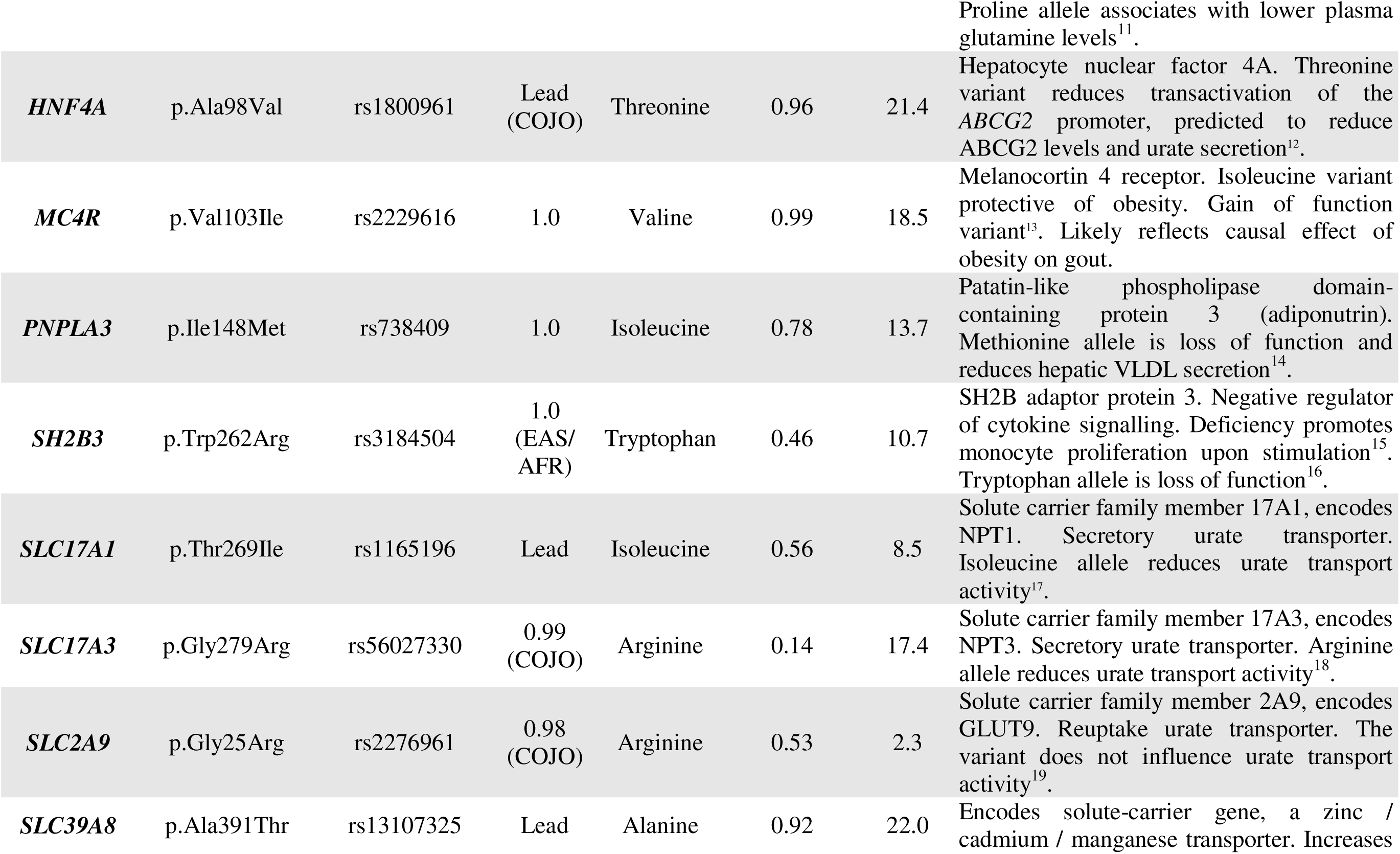
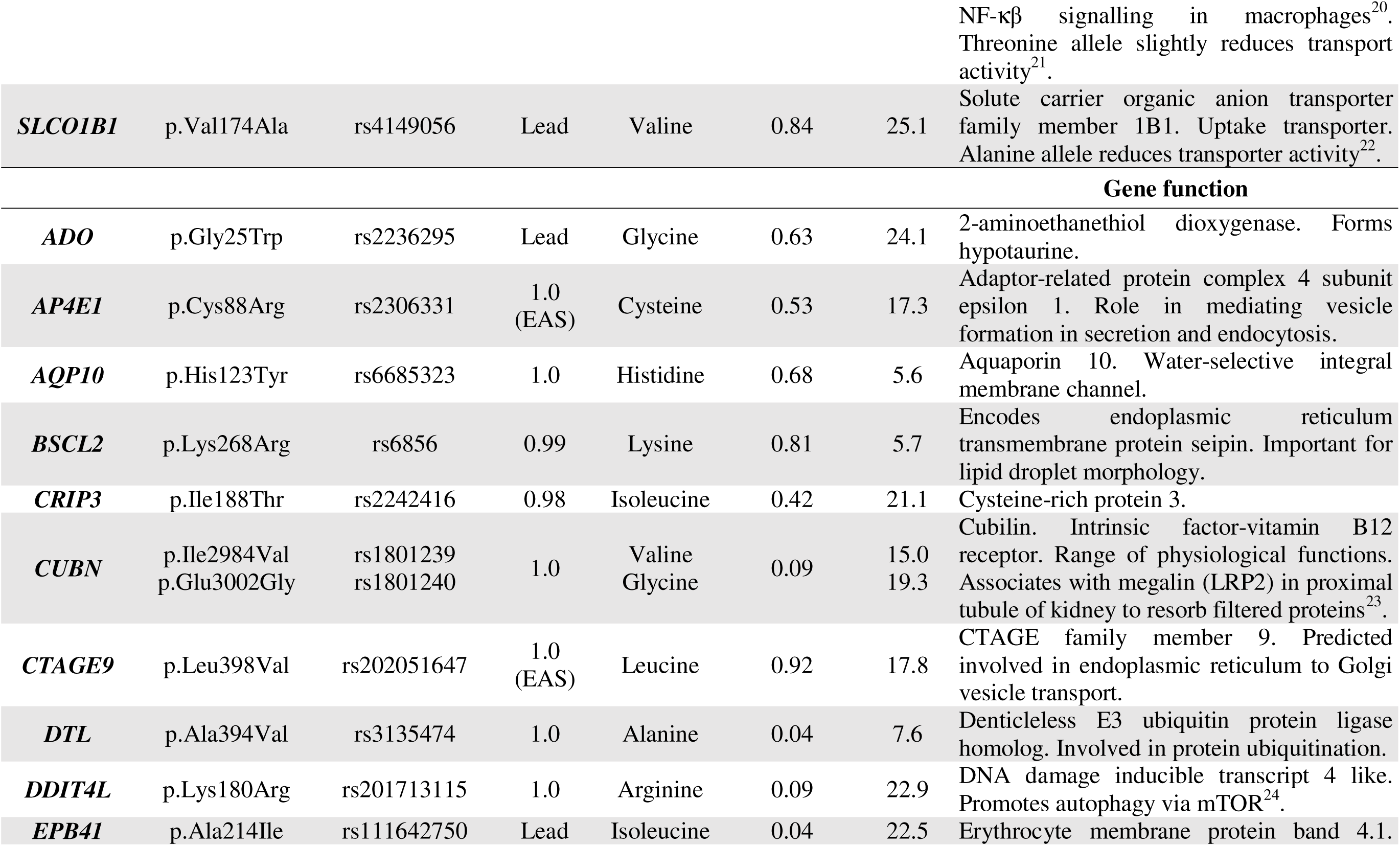
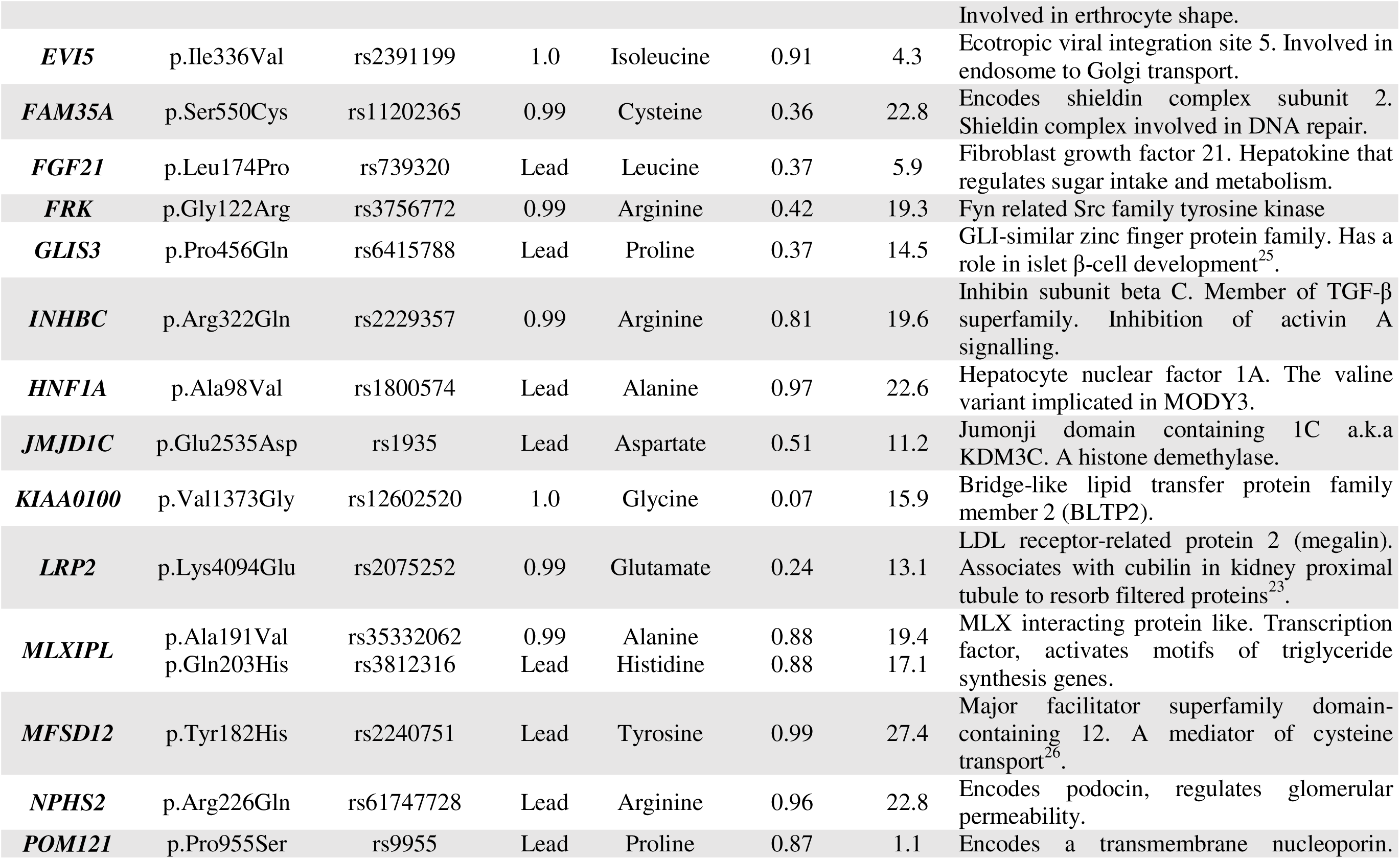
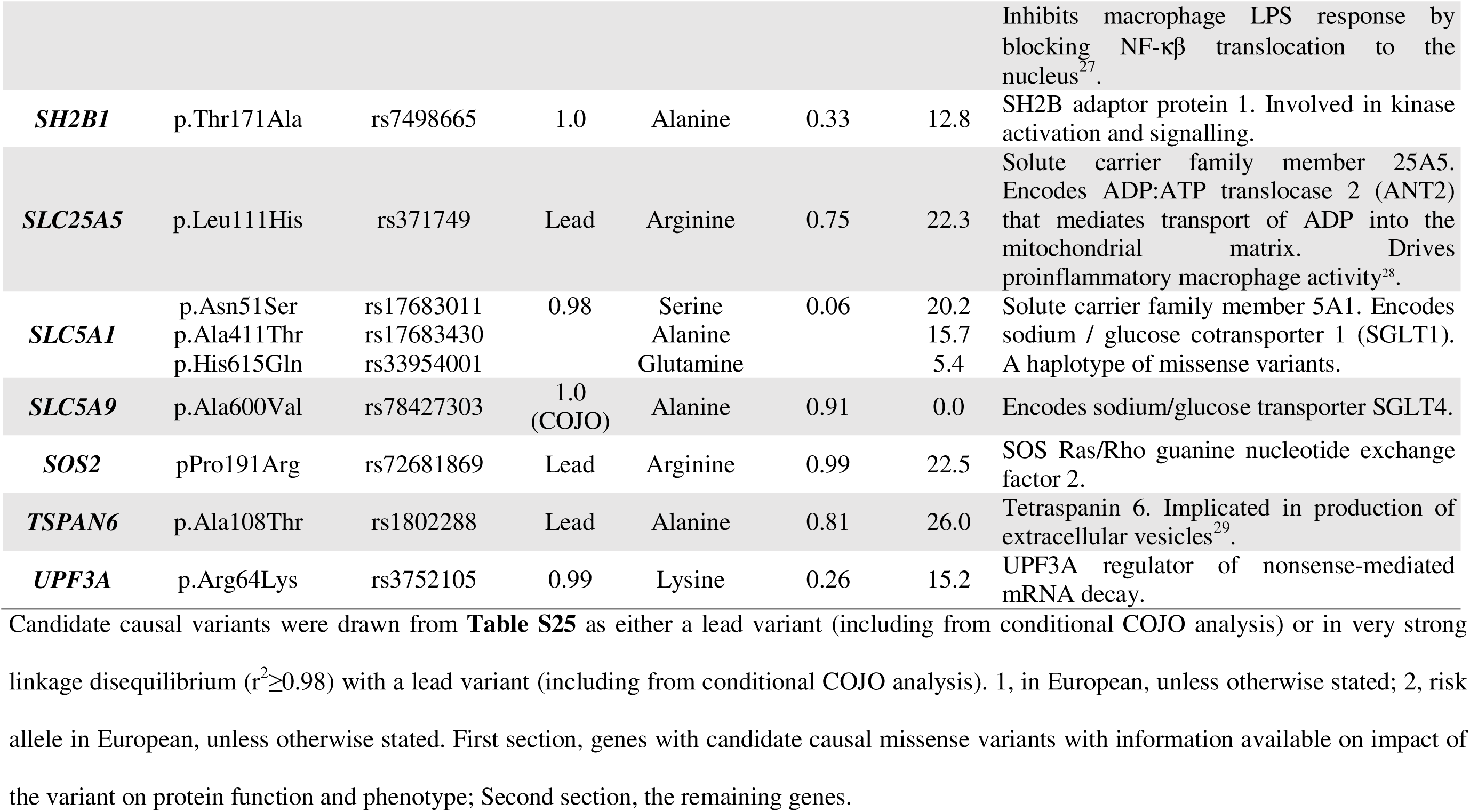
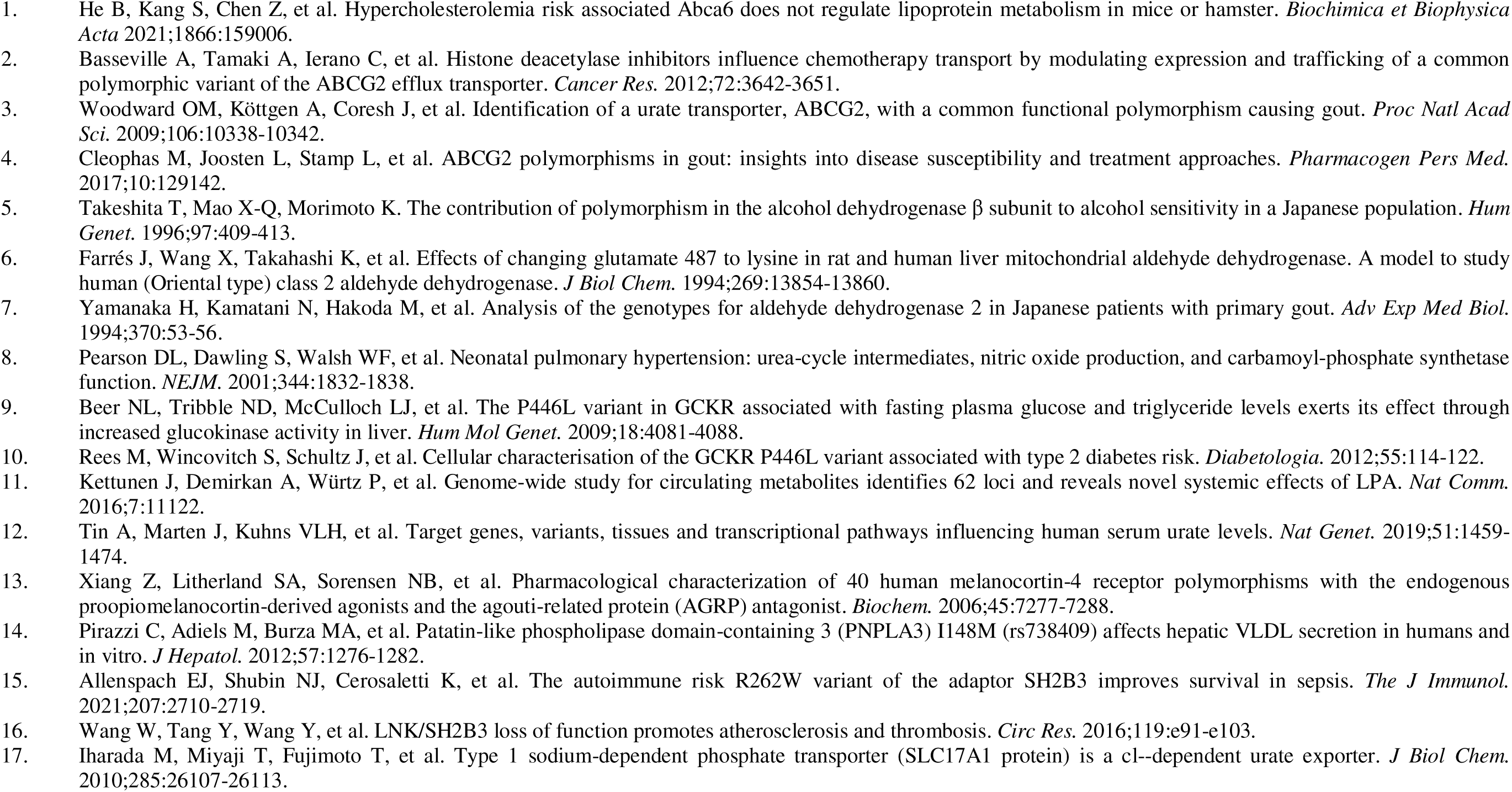
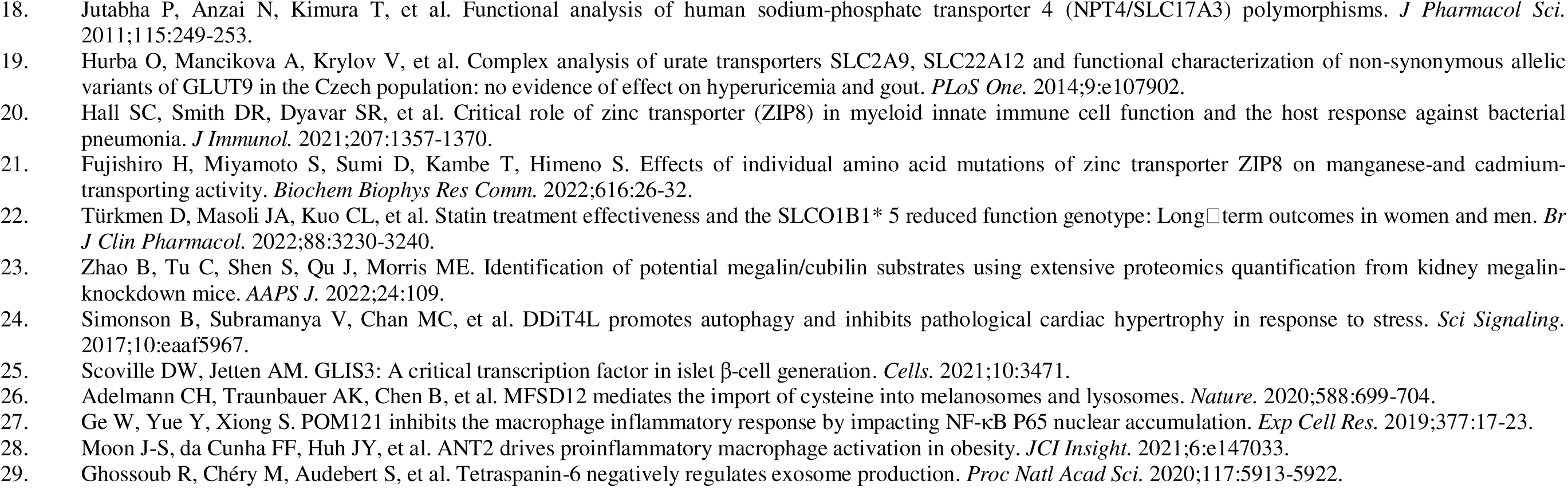
Table of missense lead variants or variants that are in high LD with lead variants. Candidate causal variants were drawn from Table S19 as either a lead variant (including from conditional COJO analysis) or in very strong LD (r^2^ ≥ 0.98) with a lead variant (including from conditional COJO analysis). 1, indicates LD in European, unless otherwise stated; 2, indicates frequency of risk allele in European, unless otherwise stated.

#### Candidate causal non-coding variants

To further investigate the remaining 1,445 non-coding candidate causal variants, we used the FATHMM^51^ non-coding score to determine if the SNP was predicted to be deleterious, and overlapped with activity-by-contact enhancer regions to determine if the variant mapped within an enhancer region that contacts a gene promoter^35^. Ninety-nine variants (7.0%) were predicted to be deleterious based on the FATHMM score for non-coding variants (FATHMM score ≥ 0.5), 256 variants (18.0%) were within an ABC-predicted enhancer for at least one gene (ABC-score ≥ 0.015), and 32 SNPs had a high FATHMM score and were also within an ABC enhancer that mapped to 78 genes (**Figure S11**, **Table S26**). Of these 32 SNPs, 29 overlapped enhancer marks and/or DNase hypersensitivity regions in blood, and ten had a co-localized eQTL for the gene(s) with which they physically interacted through an ABC enhancer (**Table S27**). Of the ten only two had eQTL in whole blood: *rs2439303* (*NRG1*) and *rs2645479* (*RPS6KB1*).

### Investigation of underlying function

#### Pathway analysis

To gain insights into biologic mechanisms we carried out pathway and gene ontology (GO) analyses using the 1,768 genes (**Table S28**) from the combination of missense genes with candidate casual missense variants (**Table 1**), co-localized eQTL genes, and the significant MAGMA genes (**Table S29**). These analyses were significantly enriched for the GO term urate metabolism, REACTOME pathways Chromatin Organization and Chromatin Modifying Enzymes, and the KEGG pathways Type II Diabetes Mellitus and PI3K-Akt signalling pathway (**Figure 4**). The 46 genes that contribute to both the REACTOME Chromatin Organization and Chromatin Modifying Enzymes terms included histone methyltransferases *EZH2, KMT2A, SETD1A, SETD2*, and three lysine demethylases *KDM3A, KDM4C*, and *KDM6B*. The 15 genes that contribute to the KEGG Type II Diabetes Mellitus and the 59 genes that contribute to the PI3K-Akt signalling pathways, include the insulin receptor (*INSR*), insulin substrate receptor 1 (*IRS1*), insulin growth factor 1 receptor (*IGF1R*), NFκΒ subunit *RELA*, p70S6 kinase (*RPS6KB1*), and *PIK3CB*.

**Figure 4:**
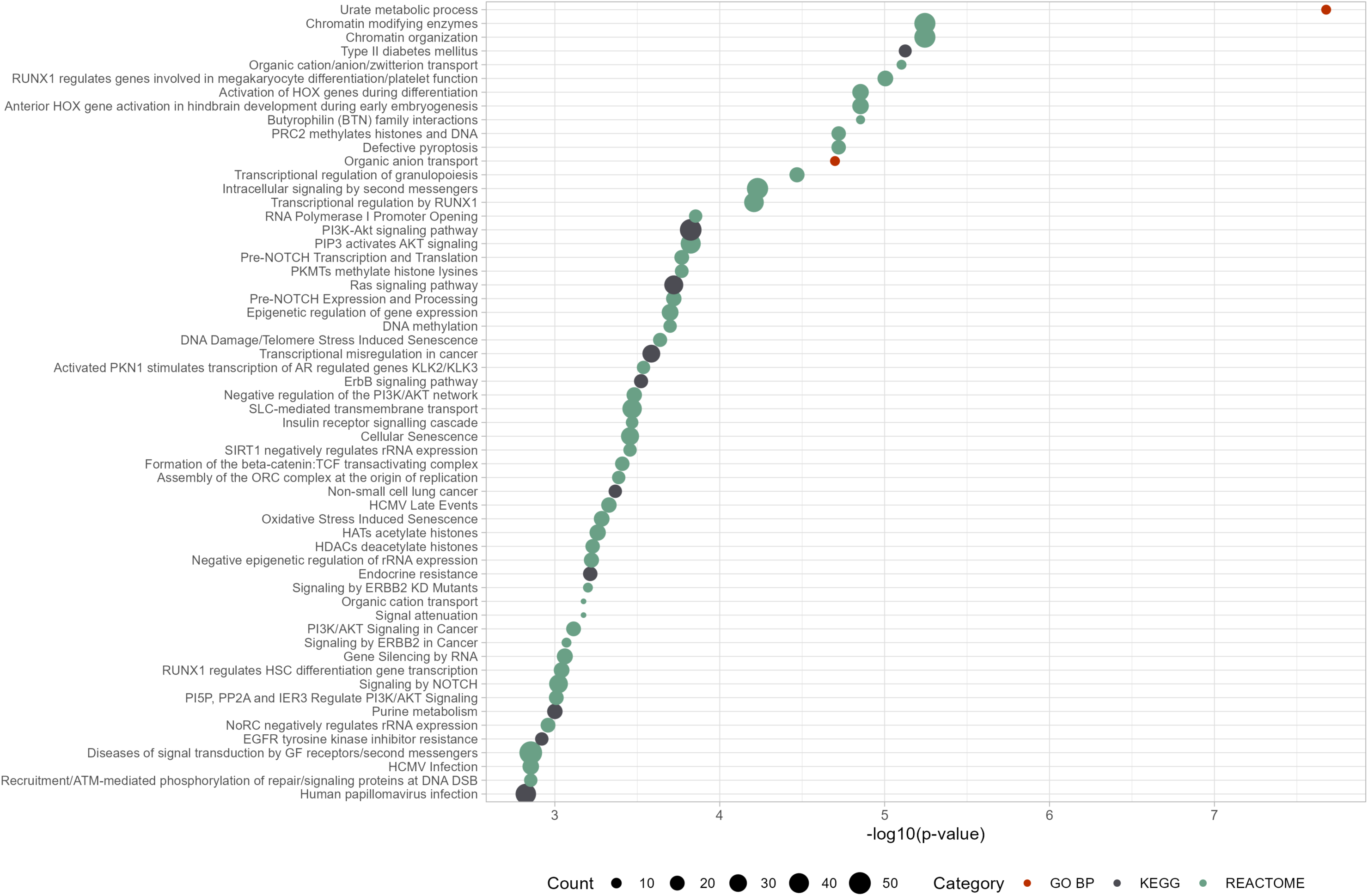
Functional and pathway enrichment analyses of gout candidate genes. The DAVID database was used to identify GO Biological Function term, KEGG, and REACTOME pathways enriched in the gout GWAS dataset. Significance (FDR) of the enrichment is denoted on the y-axis, size of the circle denotes number of genes contributing to the enrichment term.

#### Prioritization of genes predicted to control gout inflammation

To identify candidate causal genes for follow-up that are more likely to be involved in the progression from hyperuricemia to gout, we developed a set of gene-centric criteria (**Methods**) that we applied to 5,014 unique genes within the boundaries of all identified GWAS loci and an additional 412 *cis*- and *trans*-eQTL genes outside the boundaries (**Table S30**). Following application of the gene-based criteria to the 5,426 genes we sub-ranked the genes within each score by function-agnostic criteria: 1) gene with closest transcription start site; 2) implicated by activity-by-contact; and 3) containing a strong candidate missense causal variant (**Table 1**). This prioritization scheme revealed strong candidate genes for a role in the initiation and resolution of the gout flare (**Figure 5**). We found 117 genes with prioritization scores between four and seven, with transcription start sites closest to the lead gout-associated SNP and/or evidence from activity-by-contact for regulation of expression by an enhancer. This prioritization evidence strongly warrants follow-up analyses in these 117 genes. Among these genes are *FADS1, DGAT2*, and the highest scoring gene *FADS2*, that are all involved in lipid metabolism, and *IRF5, IL1R1, TRAF4, IL6R, IK,* and *MAST3* all implicated in the inflammatory response. To support our prioritization approach, we identified category enrichments using DAVID^52^ for the gene-disease association dataset class category ‘Immune’ (*P* = 2.9×10^−11^) and the transcription factor NFκB (*P* = 3.8×10^−17^) for 585 genes scored ≥ in the prioritization scoring (**Table S31**). These enrichments were maintained in the 519 genes that remained when we excluded the 66 genes at the HLA locus (*P* = 1.3×10^−3^ and 2.5×10^−15^, respectively; **Table S32**).

**Figure 5:**
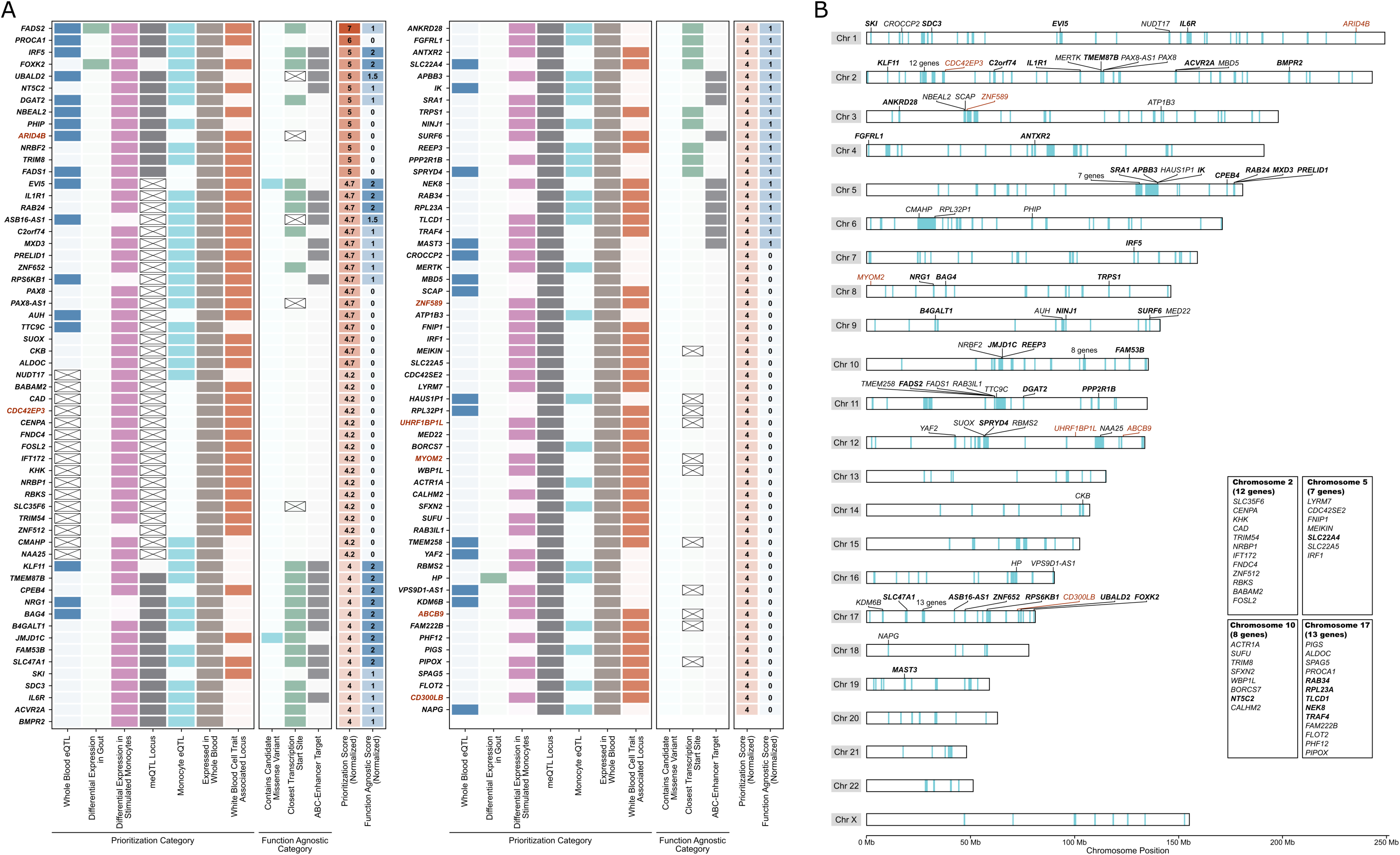
Genes prioritized for a role in gouty inflammation. **A)** 117 genes with a normalized prioritization score ≥ are listed from highest to lowest score. The seven prioritization categories (left), three function agnostic categories (middle), and the normalized scores (right) are given for each gene. Cells are colored if the gene gained a point in the prioritization/function agnostic scores based on the criteria of that category and crossed if the category data was unavailable for that gene. Red gene labels represent those that were identified as a *trans*-eQTL. **B)** ideogram showing the genomic location of the 117 genes with a prioritization score ≥ Bolded gene labels represent those that had a function agnostic score ≥1 and red gene labels represent those that were identified as a *trans*-eQTL. Light blue highlighting within the ideogram chromosomes indicates the genomic location of all significant loci identified, amalgamated across all ancestry-, trans-ancestry, and sex-specific analyses.

#### The IL1RN locus and IL-1β response to monosodium urate crystal stimulation

The Human Functional Genomics Project (500FG) is a repository of cytokine QTL (cQTL) data, including data on association between IL-1β and IL-6 response to monosodium urate crystal/C16 stimulation in peripheral blood mononuclear cells^53^. We investigated the associations between the 500FG IL-1β or IL-6 response to stimulation and 41 lead SNPs from the European or trans-ancestry GWAS that were not associated with urate in the UK Biobank (**Methods**) and had genotype data available in the 500FG dataset (either the variant itself or a proxy; r^2^ ≥ 0.8) and of 202 lead variants associated with urate. There was evidence of amplification of association signals in the 41 non-urate-associated SNPs for IL-1β response, but not in the 202 urate-associated SNPs (**Figure S12, S13**). The lead SNP at the *IL1RN* locus was the only SNP significantly associated with IL-1β response to monosodium urate crystal stimulation (*rs9973741*; risk-allele (G) β = 0.34, *P* = 3.6×10^−4^ < 0.05/41). IL-6 response to monosodium urate crystal/C16 stimulation also associated with *rs9973741* (β = 0.32, *P* = 2.4×10^−4^). This SNP did not associate with either response upon stimulation by *C. albicans* (β = 0.04, *P* = 0.20 and β = −0.07, *P* = 0.19, respectively).

Association signals at this locus co-localized between gout and genetic control of *IL1RN* expression in testis and sub-cutaneous adipose tissues, and *IL1F10* (*IL-38*) expression in skin (**Figure S14**). The G-allele associated with increased expression of *IL1RN* in the testis and skin (β = 0.29, *P* = 4.0×10^−11^; β = 0.13, *P* = 1.7×10^−8^) and reduced expression in sub-cutaneous adipose (β = −0.13, *P* = 2.0×10^−5^), and with reduced expression of *IL1F10* in the skin (β = −0.18, *P* = 1.1×10^−9^). Collectively, these findings are consistent with the hypothesis that the genetic influence on the risk of gout occurs through control of IL-1β production by the innate immune system when activated by MSU crystals, contributing to the control of *IL1RN* and/or *IL1F10* gene expression.

#### Xanthine oxidoreductase

Xanthine oxidoreductase (XOR), the sole enzymatic source of urate that oxidizes hypoxanthine to xanthine and then xanthine to urate, is a key gout intervention point with urate-lowering therapy. XOR is transcribed and translated as xanthine dehydrogenase (XDH), with the circulating form exhibiting dehydrogenase and oxidase activity. Given the key role of XOR in the pathogenesis of gout and evidence for association of a burden of rare variants in *XDH* with reduced urate level^54^, we explored the significant association of the *XDH* locus with gout in more depth. There were clear signals of association with urate in the independent UK Biobank (**Methods**) and Tin et al.^9^ studies that overlapped the gout signal (**Figure 6**), with the gout risk allele in strong LD with the urate-increasing allele in each of the urate datasets. At the *XDH* locus we identified a *cis*-eQTL for *XDH* that was specific to the prostate (**Table S15**). The gout risk allele associated with increased *XDH* expression in the prostate. There were eQTL for *XDH* in other tissues (the notable exception being the liver) but none co-localized with the gout genetic association signal (**Figure 6**). Sex-stratified analysis of the gout GWAS data provided evidence for an association signal in men but not in women (**Figure 6**; *rs7594951* C-allele OR_female_ = 1.02 [95% CI; 0.99-1.05], OR_male_ = 1.05 [95% CI; 1.03-1.08]).

**Figure 6:**
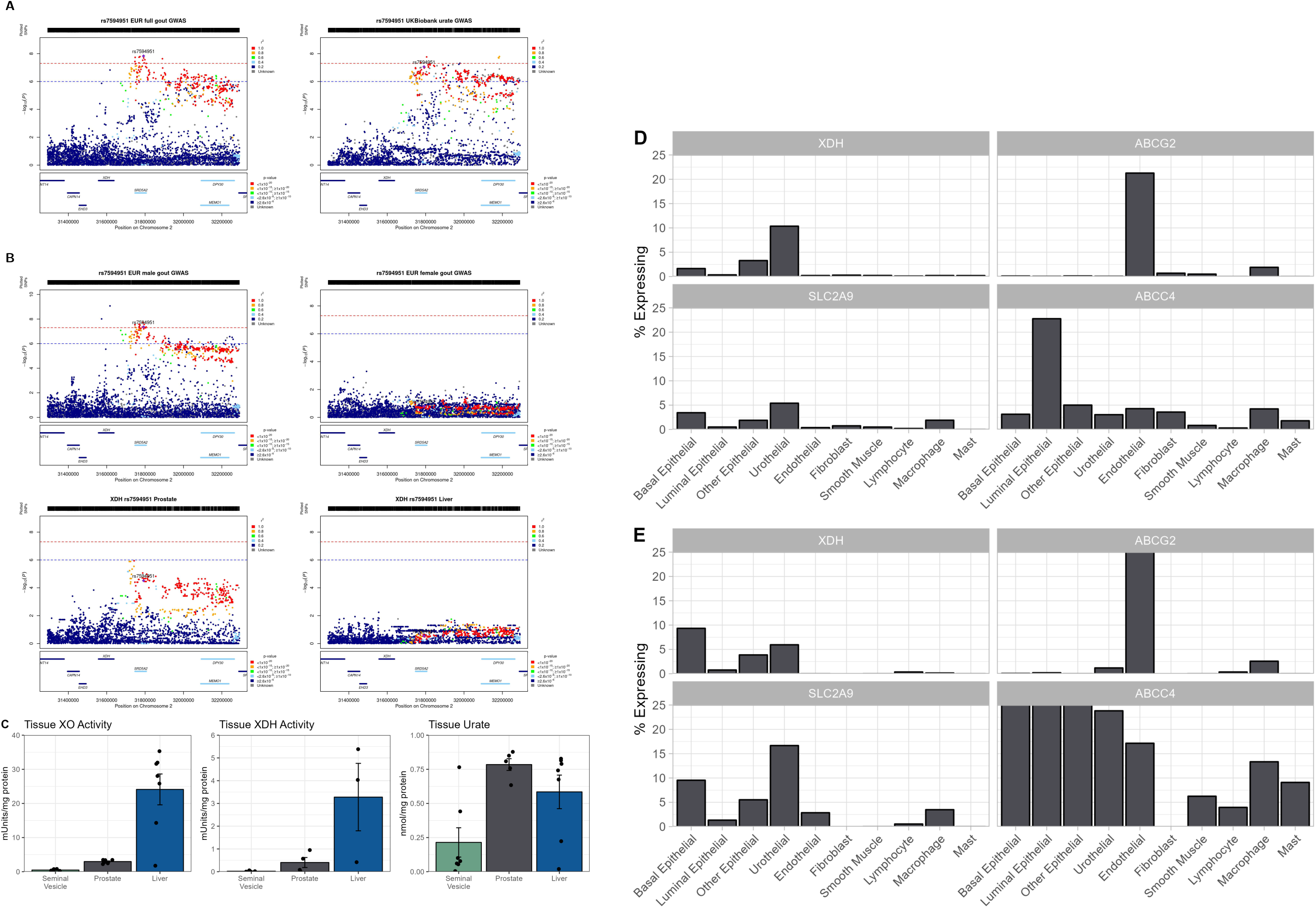
*XDH* expression and xanthine oxidoreductase (XOR) activity in the prostate. **A)** Locus Zoom plots of genetic association at the *XDH* locus with gout (European full analysis; left) and serum urate in Europeans (UK Biobank; right). Each genetic variant is represented by a single dot plotted according to genomic position on the x-axis with strength of evidence for association (-log_10_P) on the y-axis. The lead variant is highlighted with a purple diamond; other variants are colored according to their LD with the lead variant (strong (red) to weak (dark blue)). **B)** the European male gout (top left) GWAS signal co-localizes with *XDH* expression in prostate tissue (GTEx v8; bottom left), but not with the European female gout GWAS signal (top right). Shown also is the expression signal of *XDH* in the liver (GTEx v8; bottom right). **C)** XOR activity and urate levels in prostate, liver, and seminal vesicle tissue from seven-week old C57Bl/6 male mice. A Kruskal-Wallis test was used to determine whether the differences between tissues were significant. **D)** and **E)** bar plots show the percentage of cells expressing *XDH* in a given cell cluster, alongside three urate transporter genes expressed in the prostate (*ABCG2*, *SLC2A9*, *ABCC4*) in prostate cell clusters. Single cell RNA-seq data were obtained from Crowley et al.^154^ (panel D) and Henry et al.^155^ (panel E).

The presence of urate in seminal fluid,^55^ urate crystals in 19/40 (47.5%) of non-malignant prostate sections,^56^ and the expression of *XDH* in the prostate (gtex.org) suggests that the prostate synthesizes urate. Supporting this we demonstrated Xor activity and urate content in the prostate of 7-week-old C57Bl/6 mice, with the urate concentration equivalent (per mg of protein) to that in liver (**Figure 6**). Our genetic data in humans implicate shared genetic control of prostate *XDH* expression and the risk of gout in men, suggesting that the prostate may secrete urate into the blood. Investigating this possibility further, we first assessed expression of urate transporters in GTEx data. Three genes encoding urate transporters were expressed: *ABCC4* (median transcripts per million (TPM) of 1.73, tissue with second highest expression after bladder); *ABCG2* (TPM of 7.6, widely expressed); and *SLC2A9* (TPM of 3.5, tissue with 4^th^ highest expression after kidney, esophageal mucosa, bladder). There was negligible expression of *SLC22A6-A8, SLC22A11/A12,* and *SLC17A1-A4*. Analysis of prostate single-cell RNA expression data showed that the expression of both *XDH* and *SLC2A9* occurs in basal epithelial cells, *ABCC4* is expressed in basal and luminal epithelial cells and the expression of *ABCG2* is restricted to endothelial cells (**Figure 6**).

#### Clonal hematopoiesis of indeterminate potential (CHIP)

CHIP is defined as the presence of clonally expanded cells with somatic mutations in an individual with no evidence of hematologic malignancy. Mutations are present in a suite of 71 genes, most prominently in genes that encode epigenetic modifiers. Recently CHIP has been associated with incident^57^ and prevalent gout^58^ but not urate^57^ and has been hypothesized to play a role in epigenetic reprogramming of the innate immune system to be more responsive to stimulation by monosodium urate crystals^59^. Given also that our pathway analysis also revealed chromatin modification to be a significant enrichment term, we investigated if germline genetic variation in the loci harbouring the 71 CHIP genes could play a role in gout. Nineteen of the 71 genes (**Table S33**) were present in the 2,598 protein-coding genes (out of a total of 21,474 from GenCode) within the boundaries of all identified gout-associated loci, a significant over-representation (*P* = 0.0002), consistent with the hypothesis that the CHIP pathway plays a causal role in gout. Enabled by a GWAS for CHIP risk alleles,^57^ we performed two-sample Mendelian randomization to test for a causal relationship of the CHIP phenomenon for gout (**Figure 7; Table S34**). There was evidence for a positive causal role of *DNMT3A*-CHIP in gout (weighted median MR estimate 0.034, *P* = 0.003; inverse variance weighted MR estimate 0.061, *P* < 0.001) but not for CHIP *per se* or *TET2*-CHIP. For the *DNMT3A*-CHIP analysis, there was no suggestion of horizontal pleiotropy by MR-EGGER (intercept 0.013, *P* = 0.23; **Table S34**).

**Figure 7:**
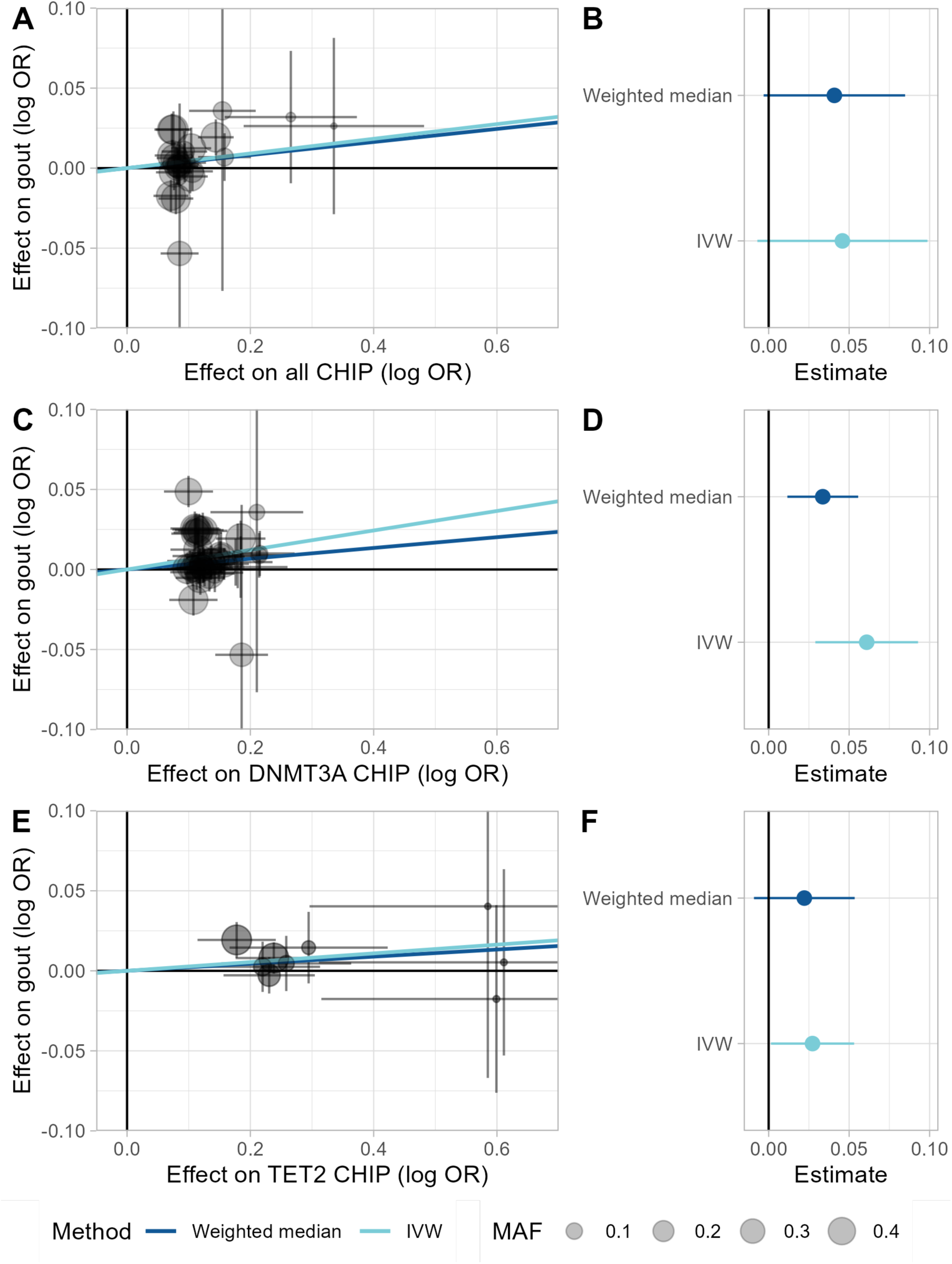
Mendelian randomization of CHIP vs gout for all CHIP, *DNMT3A* CHIP, and *TET2* CHIP. **A)**, **C)**, and **E)** are plots of the relative effect sizes of CHIP-associated variants on gout for each of the three CHIP types, respectively. Overlaid lines indicate the Mendelian randomization estimates from weighted median, inverse variance weighted, or MR-Egger regression. **B)**, **D)**, and **F)** show the Mendelian randomization estimates as forest plots for each of the three CHIP types, respectively, including the MR-Egger intercept for each model.

#### Loci with cis-eQTL for protein-coding and lncRNA genes – candidate immune-priming lncRNAs

Of the lead variants, 212 were a *cis*-eQTL for a protein-coding gene, of which 75 (35.3%) also had a *cis*-eQTL for a long non-coding (lnc) RNA. The lncRNAs at these 75 loci are candidate immune-priming lncRNAs (IPLs) that may prime immune genes for robust transcription by facilitating transcriptional machinery assembly (e.g. the prototypical UMLILO lncRNA^60^), and can be regarded as potential targets for RNA-mediated therapeutics. Indeed, two gout associated loci containing *CSF1* and *IRF1* were previously identified as being connected by a IPL^60^. To identify candidate IPLs amenable for follow-up mechanistic study, we prioritized the 75 loci based on the protein-coding gene having an eQTL in whole blood, and a high prioritization score (**Table S30**) for a role in the progression from hyperuricemia to gout (**Table 2**). We also included in this set the lncRNA DRAIR, previously functionally implicated in the immune response of macrophages^61^. Two of the eQTL-lncRNA pairs both had an eQTL in whole blood (*DGAT2/RP11-535A19.2* and *BAG4/RP11-350N15.5*).

**Table 2:**
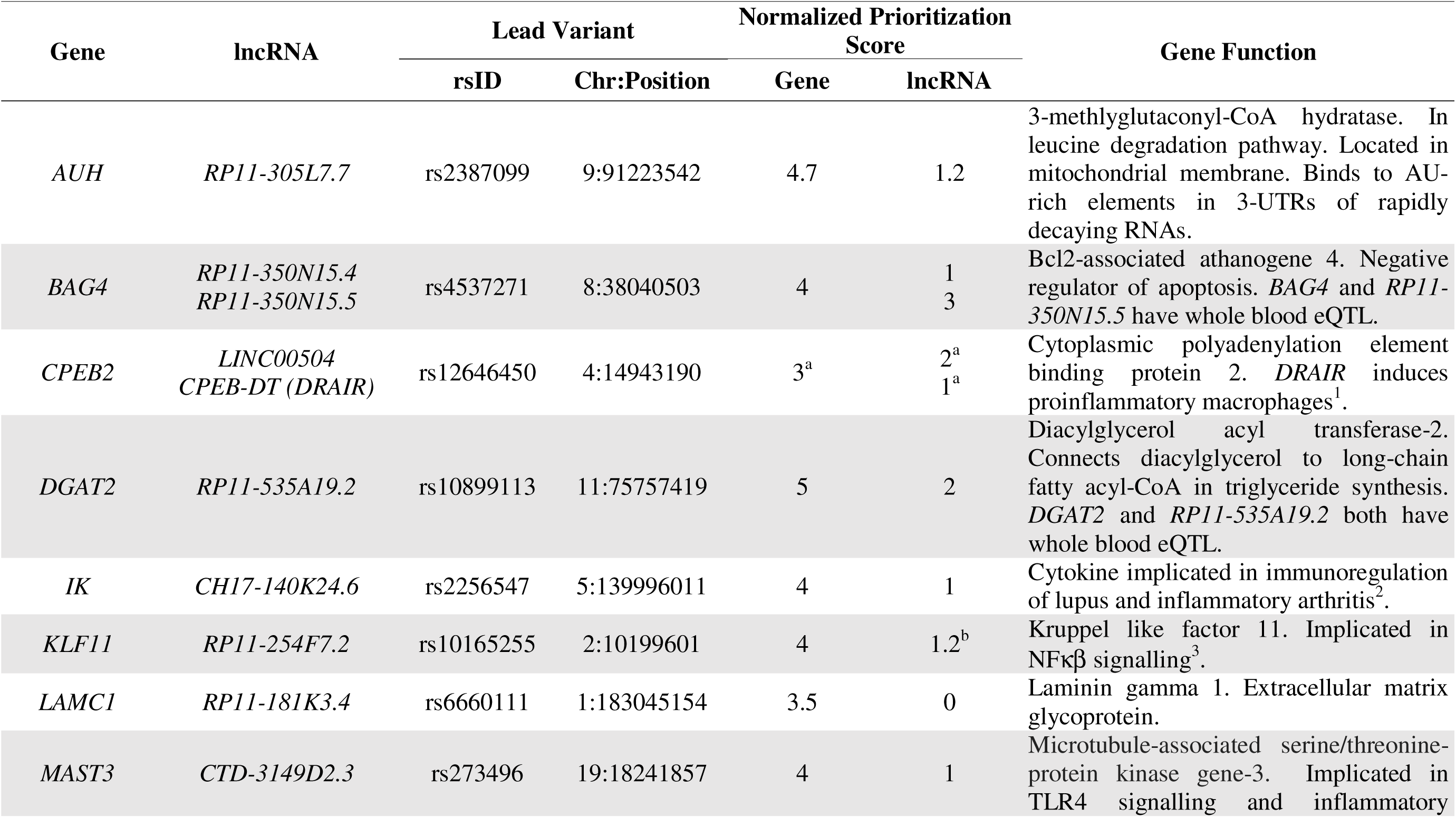
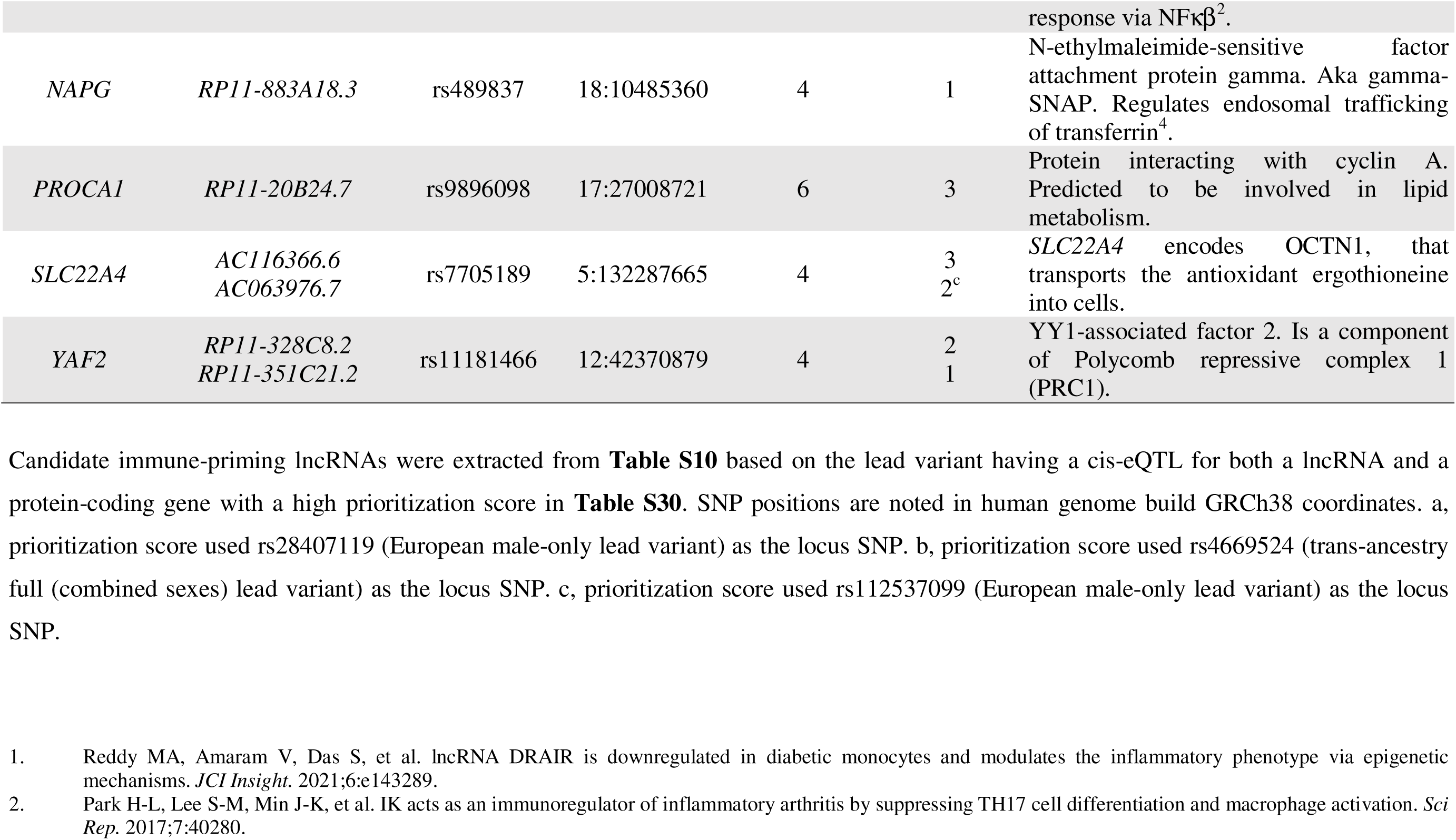
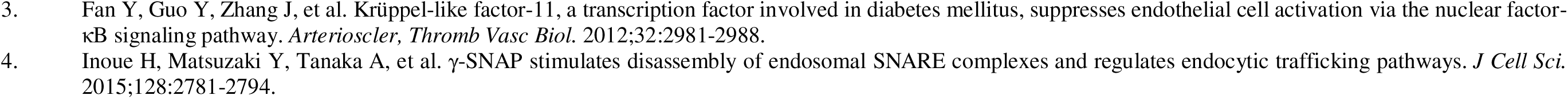
Table of candidate immune-priming lncRNAs. Candidate immune-priming lncRNAs were extracted from **Table 120** based on the lead variant having a cis-eQTL for both a lncRNA and a protein-coding gene, and the gene having a high prioritization score in **Table S30**. SNP positions are noted in human genome build GRCh38 coordinates. a, prioritization score used *rs28407119* (European male-only lead variant) as the locus SNP. b, prioritization score used *rs4669524* (trans-ancestry full (combined sexes) lead variant) as the locus SNP. c, prioritization score used *rs112537099* (European male-only lead variant) as the locus SNP.

We used GeneHancer to examine evidence of physical connections between the lead genetic variants, the lncRNA and the protein-coding gene at *DGAT2* (**Figure 8**) and other genes in **Table 2** (**Figure S15**). We identified physical connections between the lncRNA and the protein-coding genes at *DGAT2* and other loci. Of particular interest is the highly prioritized gene *DGAT2* (**Figure 5**). The gout risk allele associates with decreased *DGAT2* and *RP11-535A19.2* expression. The promoters of *DGAT2, RP11-535A19.2* and *UVRAG* are marked by H3K4me3 (**Figure 8**). There are also six CpG sites at DGAT2 with co-localized meQTL. The gout risk allele associates with increased CpG methylation at all sites except cg02337499, which is found at the promoter of *UVRAG*. A GeneHancer physical connection was observed between a maximally associated variant *rs11236533*, the lncRNA and *DGAT2*. Functional data for *rs11236533* indicates that it is bound by CTCF (a CHIP gene which also maps to a male-only gout-associated locus) in numerous cell types including monocytes and neutrophils and is predicted to alter a CTCF motif. *Rs11236533* physically connects to *DGAT2* via two GeneHancer regions containing additional maximally associated variants (region 1: *rs7947512, rs10899119, rs11823869* and *rs10219339;* and region 2: *rs11236530*). In the ImmuNeXUT dataset^62^, *rs11236533* is the lead variant for an eQTL for *DGAT2* and *CTD-2530H12.2* in neutrophils and the gout risk allele decreases expression of both genes in blood. Collectively these data suggest the possibility that this region is under immune cell-specific regulatory control that may be mediated by DNA methylation and the expression of lncRNA *RP11-535A19.2* at this locus.

**Figure 8:**
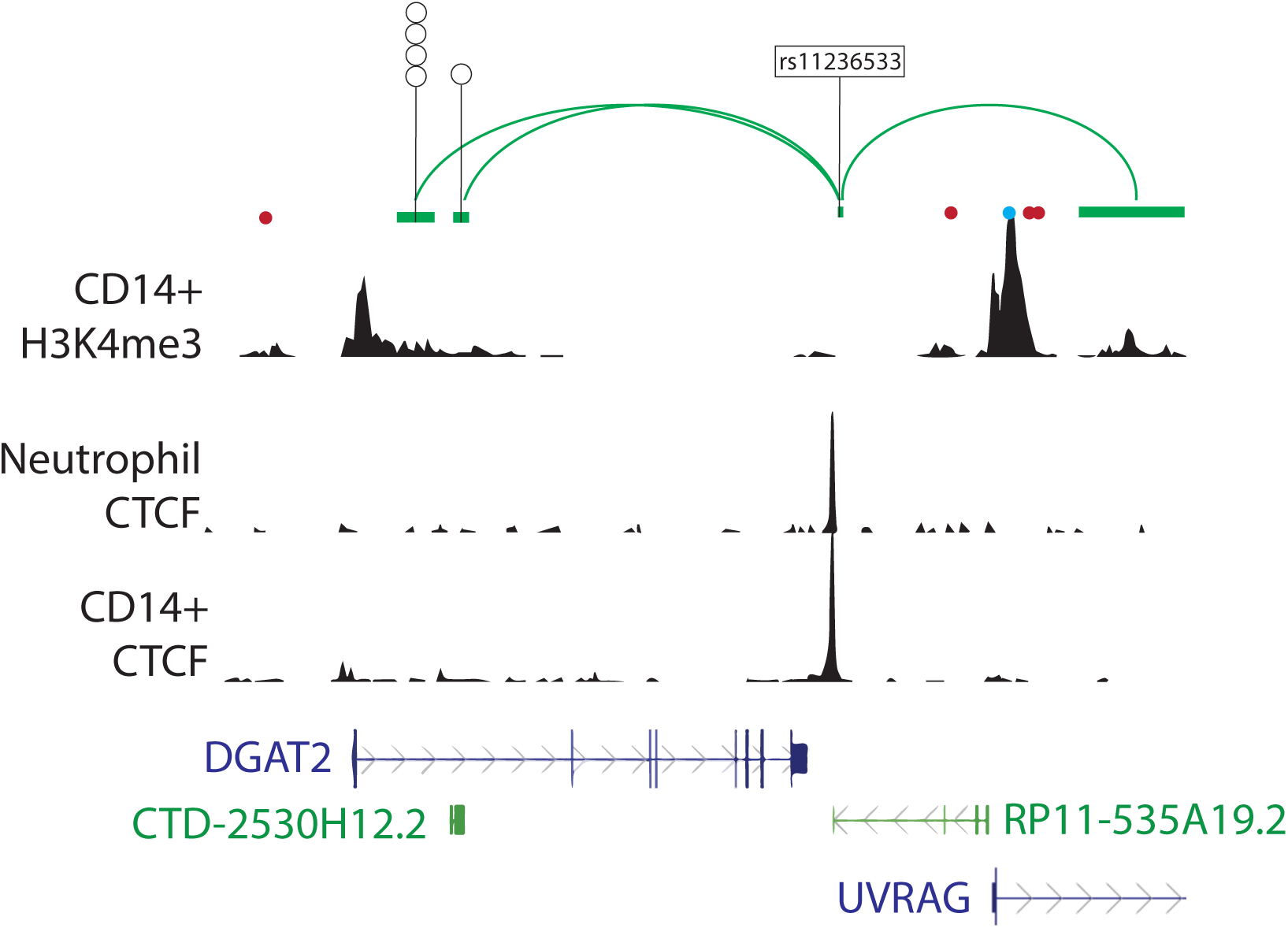
*DGAT2*: Example of genome organization at a candidate immune-priming lncRNA locus. ENCODE H3K4me3 signal track from CD14+ monocytes indicates enrichment at the promoters of *DGAT2*, the lncRNA *RP11-535A19.2,* and *UVRAG*. ENCODE CCCTC-binding factor (CTCF) signal track from neutrophils and CD14+ monocytes indicates CTCF binding at the SNP *rs11236533*. Genehancer connections are indicated in green and illustrate physical connections (Hi-C) between *rs11236533,* which disrupts a CTCF binding site and additional maximally associated SNPs at two Genehancer regulatory elements. Red and blue dots indicate the CpG locations that are associated with co-localized meQTL and the colour denotes direction of effect of the gout risk allele (red = higher methylation, blue = lower methylation).

#### Drug targets

We took a systematic approach to identifying drug targets using the Genome for Repositioning (GREP) drugs pipeline. We used MAGMA^28^ (for generalized gene set analysis of GWAS data) on the combined trans-ancestry meta-analysis to identify 476 genes (**Table S35**) for input into GREP (**Figure S16**)^63^, which examines a gene set for enrichment in drug targets by clinical indication category (Anatomical Therapeutic Chemical Classification System (ATC) or International Classification of Diseases 10 (ICD10) diagnostic code). In the ATC analysis the only category nominally significant was ‘antigout preparations’, driven by target genes *SLC22A12* (encoding URAT1) and *SLC22A11* (encoding OAT4) and uricosuric drugs lesinurad, sulfinpyrazone and probenecid. Xanthine oxidase inhibitors were excluded because the *XDH* gene was not significant (*P*=1.0) in the MAGMA analysis and was not included in the input gene list. In the ICD10 analysis nominally significant categories were neoplasms, blood biochemistry and metabolic disorders, driven by the *CASP9, CCND1, CHEK2, DRD5, IGF1R, INSR, PPARG* and *DGAT2* genes. The ICD10 M5-M14 ‘inflammatory polyarthropathies’ category that includes gout was not significant (*P* = 0.16), nor was ICD10 M10 gout code *(P*=0.59).

## Discussion

The sample size of this trans-ancestry gout GWAS (2.6M individuals) represents a 3.2-fold increase in people with gout over the previous largest study, the Global Biobank Meta-analysis Initiative that detected 52 loci^16^. The increase in sample size allowed a substantial increase in detected loci, to 376 in total (of which 148 were new in urate and gout), facilitating new findings into the molecular pathogenesis of gout. Study size allowed us to perform sex-specific GWAS and to discover 39 loci not detected in the main analysis. We present evidence for genetic control of expression of *XDH* that co-localizes with genetic association with gout and urate levels only in the prostate, implicating the prostate in urate homeostasis, and we provide evidence for a causal role of clonal hematopoiesis of indeterminate potential in gout. Top pathways identified are synthesized into **Figure 9**. Given previous major insights into the molecular control of urate levels by GWAS^9,25,64,65^, we focused this study on identifying molecular mechanisms of the progression from hyperuricemia to symptomatic gout for which we generated a comprehensive list of target genes for follow-up studies.

**Figure 9:**
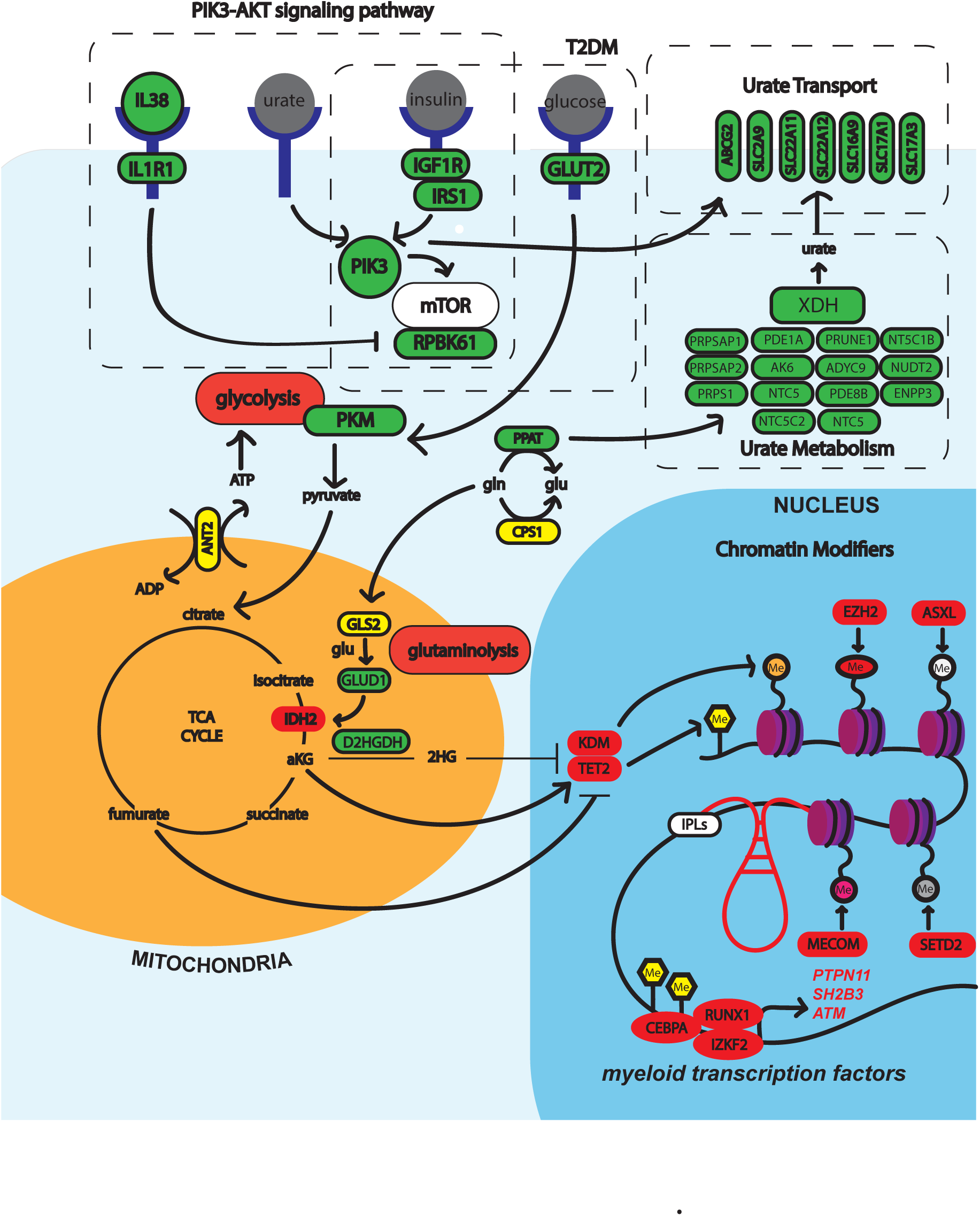
Linking of urate metabolism and transport, insulin signalling, the Krebs cycle, and epigenomic reprogramming with candidate gout-associated genes. Candidate genes are encircled in bold lines, CHIP-implicated genes in red bold lines.

Co-localization of the male gout genetic association signal at *XDH*, with genetic control of *XDH* expression only in the prostate, with the gout-risk allele associated with increased prostate expression of *XDH*, generates the hypothesis that the human prostate synthesizes urate, contributing to increased urate levels and higher risk of gout in men. The relative contribution of different tissues to circulating urate in humans is not quantified, although the liver is a substantial contributor (in mouse, ~50% of plasma urate is contributed by the liver^66^ with relative contributions of other murine tissues unknown). Urate is a constituent of human semen^55,67^and MSU crystals were present in 19 of 40 (47.5%) non-malignant prostate sections^56^, suggesting that it is common to have urate super-saturation in the prostate. The supposition that the prostate exhibits XOR activity is supported by demonstration of XOR activity and urate content in the prostate of C57Bl/J6 mice with the prostate urate concentration equivalent to the liver (**Figure 6**). *XDH, SLC2A9* and *ABCC4* are co-expressed in the urothelial cells, and the gene encoding the urate secretory transporter *ABCG2* is predominantly expressed in the endothelial cells of the human prostate. This is consistent with a model of urate reuptake by GLUT9 (encoded by *SLC2A9*) from the urethra by urothelial cells, and urate synthesis in epithelial cells by XOR. Urate could then be transported out of epithelial cells by GLUT9 and/or ABCC4 and transported from endothelial cells into the circulation by ABCG2.

From the full GWAS, 36 loci had effect sizes that differed between sexes, all with a stronger effect size in men. These are dominated by previously urate-associated loci. This is consistent with the emerging consensus that urate control and subsequent risk of gout has a stronger genetic component in men (e.g. contributing to lower fractional excretion of urate in men^68^), whereas in women, who develop gout at an older age, increased urate levels are driven by a higher burden of comorbidity^27,69,70^. Excepting the African analysis, *ABCG2* consistently exhibited heterogeneity with a stronger effect size in men, consistent with the heterogeneity observed in association with urate^9^. In contrast, *SLC2A9* has a stronger urate effect size in women^9^, but a stronger gout effect size in men, as did *A1CF* and *IGFR1*. The reason for this is unclear, but it may reflect an additional role of the loci in the progression from hyperuricemia to clinical gout. Considering *SLC2A9* and *IGFR1*, in the context of urate control, there is a differential effect of *IGFR1* genotype on urate levels between men and women in the presence of the urate-lowering genotype at *SLC2A9.* Functionally, IGF1 stimulates urate reuptake by GLUT9 (encoded by *SLC2A9*) via IGFR1^71^.We developed an approach to prioritize from the gene pool encompassed by the 376 loci genes for follow-up, predicted to control the innate immune response to MSU crystals. Our primary prioritization scheme focused, by necessity, on the gene *per se,* and did not incorporate aspects of previously proposed functionally-agnostic variant-to-gene criteria^72,73^. However, we secondarily ranked on some genomic criteria which serves to increase confidence in causal candidacy. This analysis identified several notable genes that illuminate new pathways in gout. The top-ranked gene (out of 5426) was *FADS2,* with *FADS1* ranked 13^th^. These genes, adjacent on chromosome 11, encode fatty-acid desaturases involved in the synthesis of unsaturated omega-3 fatty acids that inhibit NLRP3 inflammasome activation^74^. Increased *FADS2* expression correlates with reduced circulating arachidonic acid and increased circulating IL-1β, TNF-α, and IL-6 levels^75^. Furthermore *FADS2*-knockdown reduced innate immune response to *C. albicans* and FADS2 inhibition reduces proliferation of innate immune cells^75,76^. Consistent with a role for FADS2 in gout, the risk allele (*rs61897795* G-allele) associates with increased expression of *FADS2* in blood. The genes encoding interleukin-1 receptor 1 (*IL1R1*) and interleukin-6 receptor (*IL6R*) were ranked 15^th^ and 47^th^, respectively. They both are co-localized eQTL in monocytes, are differentially expressed in monocytes exposed to lipopolysaccharide or monosodium urate crystals^43^, their TSSs are contacted by an enhancer, and they have the closest TSS to the lead gout-associated genetic variant. IL-1R1 is antagonized by anakinra, which is an effective option for treatment of gout flares^77^. Interestingly the *IL-1R1* genetic signal, along with an intergenic signal on Chr21:40.4 - 40.5Mb (*rs4817983, rs2836884*), are the only signals to be present in both the European and trans-ancestry analyses (including sex-specific) and which have not been previously reported in gout or urate related GWAS. The Chr20 locus has been found to associate with white blood cell traits^78,79^ and other inflammatory conditions including ulcerative colitis^79^ and Takayasu’s arteritis^80^. Interleukin-6 is secreted by monosodium urate crystal/lipopolysaccharide-activated monocytes ^81^. Although neither IL-6 nor IL-6R have been rigorously evaluated as targets in the management of gout, the JAK-inhibitor baricitinib, one effect of which is to block IL-6R signalling, reduces IL-1β secretion in the response of IL-6-primed neutrophils to monosodium^82￼^, and case reports indicate efficacy for the IL-6R antagonist tocilizumab in refractory gout ^83–85^.

We identified 47 genes with candidate causal missense variants. *AQP10*, encoding aquaporin 10, ranked 296^th^ in our prioritization scheme and is a co-localized eQTL in whole blood, however the lead associated variant is also in strong linkage disequilibrium (r^2^ ≥ 0.98) with two missense variants (*rs6668968*/p.Arg15Gln and *rs6685323*/p.His123Tyr). Aquaporins allow small solutes (e.g. water, glycerol) to cross membranes. *AQP10* has low levels of expression in most tissues, with higher expression in whole blood, fallopian tube, lung, spleen and testis, and transports water in a pH-independent manner and glycerol in a pH-dependent manner, playing a role in lipolysis in adipocytes^86^. The regulation of cell volume induces ionic changes that regulate NLRP3 inflammasome activation^87^ and monosodium urate crystal-induced production of IL-1β by THP-1 monocytes is reduced with inhibition of aquaporins^88^, suggesting cellular osmolarity to be a newly implicated causal mechanism in gout. The *CUBN* and *LRP2* genes encode cubilin and megalin, large endocytic proteins expressed in the kidney proximal tubule that resorb protein from the urine. Both genes also associate with urate levels and the combined activity of cubilin and megalin may influence urate levels and the risk of gout through an influence on filtration of urate by the kidney. Interestingly, the *CUBN* locus controls expression of *SLC16A9* in *trans –* the *SLC16A9* locus itself associates with urate, gout and carnitine levels^89^ and *SLC16A9* encodes a creatine transporter in the kidney proximal tubule. *SLC5A1* encodes sodium-glucose transporter 1 (SGLT1) and is primarily expressed in the small intestine. Recently, in the context of the established urate-lowering effect of SGLT2 inhibitors^90^ (SGLT2 is expressed primarily in the kidney), Mendelian randomization using the *rs17683430* (p.Ala411Thr) variant supported SGLT1 inhibition as a therapeutic option in gout^91^. Two mitochondrial enzymes *CPS1* encoding carbamoyl phosphate synthase 1 and *GLS2* encoding glutaminase 2 have candidate missense variants, with the gout risk allele of *CPS1* associated with lower arginine levels^92^ and the gout risk allele of *GLS2* associates with reduced glutamine levels^93^. The missense variants may impact Krebs cycle metabolites and possibly contribute to gout risk via immunometabolic epigenetic reprogramming through histone modifications^94^.

Clonal hematopoiesis of indeterminate potential (CHIP) occurs as a result of white blood cell somatic mutation in a suite of genes, predominantly in *TET2* and *DNMT3A*. Consistent with a previous report of association of CHIP with gout^58^ we observed an over-representation of genes mutated in CHIP mapping to gout loci. By Mendelian randomization, using instruments comprised of genetic variants associated with CHIP *per se*, *TET2*-CHIP and *DNMT3A*-CHIP, we provided evidence for a causal role of CHIP for gout, implicating CHIP (specifically *DNMT3A*-CHIP) as a causal pathway in the pathogenesis of gout. Genes mutated in CHIP and associated with gout include epigenetic regulators (e.g. *TET2, EZH2*), damage response proteins (e.g. *PPM1D*) and metabolic enzymes (*e.g. IDH2*). *TET2, EZH2* and *IDH2* are all strong candidate genes for gout, being eQTL that co-localize with gout-associated GWAS signals. *IDH2*, encoding isocitrate dehydrogenase 2, produces α-ketoglutarate in the Krebs cycle, a source of substrates for epigenomic modification and also a co-factor for histone demethylases and the TET family of demethylases^95^. It is unclear how CHIP *per se* could cause gout. However, somatic mutation in CHIP genes associated with gout,^58^ combined with knowledge of their functional roles and that inherited genetic variants influence their expression and are associated with gout, provides further evidence for epigenomic remodelling in the pathogenesis of gout. This would include epigenomic alteration of the innate immune system by soluble urate (training)^7^. Implication of the PIK3-AKT-mediated insulin signalling pathway, that may stimulate glycolysis and provide substrate for the Krebs cycle, suggests that the pathogenesis of gout involves insulin-stimulated production of substrate for epigenomic remodelling. Indeed, IGF1R and IRS1 are central to insulin signalling, are co-localized eQTLs, and have been implicated in the training mechanism^96^. CHIP also associates with heart failure^97–99^ and deteriorating kidney function^100^ and may link the etiologies of gout and co-morbid cardiorenal conditions via an inflammatory state. Supporting a role for the NLRP3-inflammasome, *TET2*-CHIP has been implicated in heart failure through the NLRP3-inflammasome and IL-1β^101^. *Tet2*-deficient mice have increased IL-1β response to MSU crystals, with *Tet2*-deficient isolated macrophages having^58^ increased IL-1β response to MSU crystal/lipopolysaccharide stimulation.

Testing lead genetic variants for association with increased IL-1β production by PBMC *ex vivo* after stimulation with MSU crystals directly implicated *IL1RN* and *IL-*38, emphasizing the IL-1β pathway as crucial in gout. The genetic signal co-localizes with genetic control of expression of *IL1RN* (encoding IL-1RA, an IL-1 receptor antagonist) and *IL38,* suggesting that the genetic influence on risk of gout is mediated by the control of expression of either or both genes. The G-allele associates with reduced expression of *IL1RN* in *ex vivo* PBMC upon stimulation with MSU crystal and C16.0 and lower levels of circulating IL1RA^102^. Consistent with these data, IL1RA acts as an inhibitor of IL-1β action by competitive binding to the IL1R1 receptor.^103^ This generates the hypothesis that IL1RA insufficiently compensates for IL-1β induction and activity in gout^103^. We did not detect genome-wide evidence for association at any genes encoding well-characterized components of the NLRP3 inflammasome or accessory molecules, nor innate molecules such as toll-like receptors, with the *IL1RN* and *IL1R1* loci representing the most well-characterized molecules that can be directly connected to IL-1β. Rather, the new genes and pathways discovered in this study (e.g. epigenomic mechanisms) likely revolve around priming and regulation of the NLRP3 inflammasome response to monosodium urate crystals. In addition to CHIP-related genes, *FADS2* and *AQP10* discussed above, other genes highly ranked in the prioritization include *TMEM176B* (encoding a transmembrane calcium channel that inhibits NLRP3-inflammasome activation)^104^, *SCAP* (encoding SREBP cleavage-activating protein, the SCAP-SREBP2 complex activates the inflammasome)^105^, and *INSIG2* (encoding a binding partner of SCAP)^106^.

A major limitation of our study is the dominance of participants of European ancestry (84.1%), with 9.2% of Latinx ancestry and only 3.6% and 3.1% for East Asian and African ancestry, respectively. This means that the new insights into the pathogenesis of gout presented here, and new interventions based on these insights, are not necessarily transferrable even to the included Latinx, East Asian and African ancestry populations, or to other non-European populations^107–109^, for example Indigenous populations in the Asia-Pacific region which suffer a high prevalence of and severe impact from gout^1,110^. There is a crisis of inequitable participation in genomics studies^107,108^. Factors causing this situation include decades of inertia from Euro-centric researchers, funders and institutions, and deep-rooted mistrust of western institutions and researchers by minority and Indigenous populations as a result of colonization, misuse of samples and deficit framing. Increasing participation of non-European participants requires well-considered addressing of concerns, meaningful engagement with and maintenance of ongoing relationships with communities for the lifetime of the research, from initial collaboration and consultation, through recruitment and the research process to when findings are generated and new knowledge is returned to communities. It is most critical to build capability of genomics researchers from under-represented populations, and to establish data governance and protection that balances the interest of communities over that of researchers^111^. The extant inequity contributes not only to under-powered GWAS but also to the lack of adequate publicly available essential genetic/genomic resources required for translation of genetic signals of association into mechanistic knowledge that include large genome sequence reference panels, body-wide eQTL datasets, and epigenomic datasets. Concerted international efforts are required to pool extant non-European genetics and genomics datasets to conduct GWAS and follow-up analyses equivalent to those done for Europeans.

Other limitations represent inherent current limitations of the field. For example, we performed co-localization analysis for eQTLs using tissues largely from GTEx and meQTL datasets generated from whole blood and kidney. With respect to meQTL in particular, additional datasets are needed across the gout disease state, for example the gut (important in urate excretion), the liver (urate production), the synovial lining (urate transport into the synovium), the kidney in different developmental stages, white blood cells and their subsets during the gout flare (e.g. monocytes) and, *ex vivo*, monocyte response to urate crystals. While we did not include replication in our study design, high correlation between gout and urate effect sizes reduces our concern about false positives. Finally, some methods of gout diagnosis (self-report and use of administrative medication data) are not as direct as clinical diagnosis. However, we have previously reported that these survey definitions of gout contributing to the Global Genetics of Gout Consortium (one of the contributing datasets (**Table S1**)) have 82% sensitivity and 72% specificity^112^, similar to a definition from the Study for Updated Gout Classification Criteria (SUGAR) that used five weighted items that included a clinical measure (hyperuricemia)^112^.

To summarize, our study reveals a broad array of strong candidate genes and molecular processes in the pathogenesis of gout suitable for follow-up studies, for example implicating the prostate as contributing to hyperuricemia in men. In particular, we have provided new insights into the molecular mechanism of the acute inflammation in gout. Our findings, including implicating the CHIP phenomenon and other epigenetic regulators, further support epigenetic reprogramming (training) as a causal mechanism in gout.

## Methods

### Study participants and phenotype definition

Study participants were sourced from 13 cohorts^11,113–121^ (**Supplementary Note, Table S1**). Gout was defined according to self-reported diagnosis and/or use of urate-lowering medication, clinical diagnosis, and/or the American College of Rheumatology (ACR) gout classification criteria^122,123^ dependent on the information available in each source cohort. Participants from these cohorts were divided into four ancestral groups (African: AFR, East Asian: EAS, European: EUR, and Latinx: LAT) based on their self-reported ethnicity or genetic ancestry. This resulted in two African, five East Asian, ten European, and two Latinx case-control study sets for analysis. Written informed consent was acquired from all participants and each study had ethical approval from the appropriate local ethics committee. One key cohort was the UK Biobank^124^ that was also used for genetic risk prediction, genetic correlation analysis with other phenotypes, Mendelian randomization and fine-mapping. 23andMe cohort participants^125^ provided informed consent and participated in the research online, under a protocol approved by the external AAHRPP-accredited IRB, Ethical & Independent Review Services (E&I Review). Participants were included in the analysis on the basis of consent status as checked at the time data analyses were initiated. Ethical & Independent Review Services was recently acquired, and its new name as of July 2022 is Salus IRB (https://www.versiticlinicaltrials.org/salusirb).

### Genotyping and imputation

Genotyping was performed separately for each of the 13 source cohorts. Cohort-specific genotyping and imputation platforms and post-imputation quality control filters are detailed in **Table S1**. Genotypes were imputed to one of the following reference panels: UK10K^126^ + 1000 Genomes Project Phase 3 (all ancestries)^127^, Haplotype Reference Consortium panel v1.1^128^, 1000 Genomes Phase 3 (all ancestries), combined Haplotype Reference Consortium panel v1.1 and UK10K + 1000 Genomes Project Phase 3 (all ancestries), HapMap phase 2^129^ (build 36), or population-specific Sequencing Initiative Suomi project v3 (build 38)^130^. The genome coordinates of the build 36 and build 38 imputed genotypes were converted to their equivalent build 37 (hg19) coordinates using snptracker v1.0 (released Dec 2014)^131^ or GATK Liftover^132^, respectively. Post-imputation quality control filters per cohort included filters for Hardy-Weinberg equilibrium, genotype missingness, minor allele frequency, and imputation quality.

### Study-specific association analysis

A genome-wide logistic regression analysis was conducted for each of the 19 case-control study sets separately with gout as a binary outcome. The regression model was adjusted for sex, age, genetic principal component vectors, and any necessary cohort-specific variables (e.g., genotyping platform) (**Table S1**). Regression analyses were repeated in male-only and female-only sub-sets for each case-control study set where possible, including the same adjusting variables except sex. Quality control steps applied to the per-study regression summary statistics included the removal of multi-allelic variants within and between study sets, exclusion of results for SNPs present in less than 5% of participants, and removal of any results outside the statistical bounds of each output (e.g. standard error = infinity). Variants with a minor allele frequency less than 0.1% or an absolute effect size (logOR) greater than 10 were also removed from the study-specific summary statistics.

### Ancestry-specific and trans-ancestry meta-analysis

An inverse-variance weighted meta-analysis of the regression results for the full, male-only, and female-only subsets was conducted for each of the four ancestral groups using METAL^133^. LD score regression analysis revealed no evidence of inflation due to factors such as population structure, therefore no genomic control correction was applied during this meta-analysis. SNPs with a heterogeneity I^2^ statistic >95% were excluded from any further analysis. Variants analysed in <50% of the case-control study sets contributing to the meta-analysis (where there were more than two contributing cohorts) and/or variants that were analysed in <20% of all contributing samples were also excluded from any further analysis.

The ancestry-specific meta-analysis results were then used as an input for trans-ancestry meta-analysis with MANTRAv1^134^. Only SNPs with an effect size present in all four ancestry groups were included in the trans-ancestry meta-analysis. Genetic variants with a meta-analysis *P* < 5×10^−8^ or log_10_Bayes’ factor ≥ 6.0 were considered statistically significant in the ancestry-specific and trans-ancestry meta-analyses, respectively (**Supplementary Notes**). Within the trans-ancestry meta-analysis, MANTRA was also used to calculate the posterior probability of heterogeneity which estimates the homogeneity in allelic effects across ancestral groups. A posterior probability > 0.90 indicates strong evidence for heterogeneity. In addition, loci with heterogenous effect sizes between sexes were identified by performing an inverse-variance weighted meta-analysis of the male-only and female-only ancestry-specific GWAS or trans-ancestry meta-analysis effect estimates. A SNP was considered significantly heterogeneous between sexes if the heterogeneity Q-statistic had a *P* < 1×10^−6^.

### Identification of genome-wide significant signals and loci

All nominally significant SNPs were identified (*P* < 1×10^−7^ for ancestry-specific GWAS or log_10_Bayes factor > 5.0 for the trans-ancestry meta-analysis) and a ±50kb window of padding was added to the SNP chromosome position. The genomic overlap of these padded chromosome positions was then identified and the outer boundaries of these regions were created using GenomicRanges^135^ v3.12. After defining these loci boundaries, any regions with ≤ 1 SNP classified as genome-wide significant (*P* < 5×10^−8^ or log_10_Bayes factor > 6.0^136^) were disregarded in further loci definitions or analyses due to potential unreliability of single variant associations. Lead SNP(s) within each of the significant loci defined above were identified by conducting LD-based clumping using PLINKv1.9b4^137^ with an ancestry-matched 1000 Genomes Project reference panel (**Supplementary Notes**). Significant loci were designated based on their chromosome and start/end position (in Mb) of the significant region (e.g. chr 5:129.52MB-131.88MB).

A regional association plot (Locus Zoom) was created for each locus (**Figure S2**) using an R script “LocusZoom-like Plots”^138^ with GENCODE human genome build 37 (release 38) gene annotation file to define gene start/end positions and gene names (HGNC gene symbol)^139^. Gene track information was coloured based on MAGMA^28^ gene-based association analysis *P*-values (Supplementary Notes). After creating these regional association plots, each locus with multiple lead SNPs was visually inspected to determine whether the labelled SNPs represented a single association signal or multiple independent association signals. For the *SLC2A9* and *ABCG2* loci with an extremely strong association signal, only a single lead SNP was included to represent the locus even if multiple lead SNPs were identified by LD-based clumping (**Supplementary Notes**). Details of all excluded lead SNPs can be found in **Supplementary Notes**. To identify additional associations, the European GWAS loci were analyzed with GCTA-COJO^140^ using the UK Biobank cohort as a reference panel (**Supplementary Notes**). Due to a lack of adequately sized reference panels for the other ancestries, we did not conduct conditional analysis on the remaining GWAS datasets.

### Previously unreported genome-wide significant loci and signals

To identify newly detected loci and signals we searched the literature for all previously reported SNPs associated with serum urate or gout. A search of GWAS Catalog^141^ was performed on 19th July 2022, using the keywords “urate”, “uric acid”, “hyperuric[a]emia”, and “gout.” This identified 68 studies published over the period of November 2007 to November 2021. All significant SNPs identified in these 68 studies were downloaded. Using build 38 genomic locations (our locus boundaries were converted to build 38 genomic locations using LiftOver^132^), we identified any previous GWAS signal that fell within our significant loci boundaries defined above. LD between the lead SNP and all previously identified SNPs in the locus was calculated using 1000 Genomes reference panels in LDlinkR^142^ with ancestry-matching to the lead SNP. LD values were categorized into high (r^2^ ≥ 0.8), moderate (0.8 > r ≥ 0.5), low (0.5 > r ≥ 0.1), or no LD (r < 0.1) groups to assess how similar the previously reported association signal was to the lead SNP. A locus was only considered new if none of the 68 urate and gout GWAS had previously reported a significant SNP within the locus boundaries.

### Testing of gout-associated loci for association with urate

332,370 unrelated participants of European ancestry were selected from the UK Biobank using self-reported and genetic (PC-determined) ancestry information (**Supplementary Notes**). Participants with evidence of gout (n = 7,131) based on hospital-diagnosed gout, self-reported gout, and use of urate-lowering medication, and those with no serum urate measurement (n = 15,531) were excluded, resulting in 309,708 participants in the association analysis for serum urate level. Variants with INFO < 0.3, MAF < 0.0001 and HWE *P* < 1.0×10^−6^ were removed. Association analysis was carried out using PLINK v1.9b6.10 with hard-called imputed genotype data, adjusted by age, sex, and PC1-40.

### Genetically predicted risk of gout

A polygenic risk score was calculated from 289 lead variants from the European GWAS that were available in the UK Biobank. The SNPs were weighted by their effect size. The resulting PRS score was divided into 10 bins. Generalized linear modeling was used to regress the PRS score against gout, adjusted by age and sex, in 332,346 European participants from the UK Biobank. The lowest PRS score bins were excluded due to an absence of gout cases. This was repeated with 316 lead variants from the trans-ancestry meta-analysis in the same cohort. Sex-specific analysis was also undertaken for each of the PRS scores. For the female analysis, the top PRS bins were also excluded. Area under the receiver operating characteristic curve (AUROC) estimates were obtained using pROC^143^.

### LD score regression with UK Biobank traits and diseases

LD score regression^29^ was used to run genetic correlation analysis of UK Biobank phenotypes with European gout GWAS summary statistics. Summary statistics for 934 primary phenotypes (**Supplementary Notes**) and baseline v1.1 reference LD scores for European ancestry was used. Since we included the FinnGen cohort in our European GWAS meta-analysis, we excluded all FinnGen phenotypes to avoid complete sample overlap (Supplementary Notes). We considered any genetic correlation with our European gout GWAS to be significant if the p-value was less than a Bonferroni-corrected significance threshold of *P* < 5.4×10^−5^ (0.05/934). The resulting significantly correlated GWAS were then tested for horizontal pleiotropy using two-sample Mendelian randomization package TwoSampleMR^144^.

### Covariate-adjusted LD score regression

LD score intercepts, cell-type group (CTG), and cell-type specific (CTS) enrichment analysis for each ancestry (African, East Asian, European, and Latinx) were determined using covariate-adjusted LD score regression (cov-ldsc)^29^. Baseline LD scores for each ancestry were generated using ancestry-specific PCs from the 1000 Genomes Project (Supplementary Notes), annotations from baseline v1.1, and LD window set to 20cM. Baseline LD scores for CTG and CTS analyses were generated similarly using the CTG and CTS annotations. The significance threshold for the coefficient p-value for the CTG analysis was set at 0.005 for a standard cut-off (for 10 cell-type groups) and 0.00125 for a conservative cut-off (for 10 cell-type groups and four ancestries), and the threshold for CTS analysis was set at 2.3×10^−4^ for a standard cut-off (for 220 cell-types) and 5.7×10^−5^ for a conservative cut-off (for 220 cell-types and four ancestries).

### Statistical fine-mapping and credible set construction

European loci were fine-mapped using PAINTOR^145^, FINEMAP^46^, and the Bayes’ factor (BF) approach^134,146^. The region to be fine-mapped was set to the locus boundary described in **Table S2** unless the region was larger than 1 Mb; in such cases the fine-mapping region was defined as the 1 Mb region around the lead SNP, but in such a way that the region to be fine-mapped did not exceed the locus boundary if the lead SNP was within 500 kb of the boundary **(Supplementary Notes**). For PAINTOR and FINEMAP, LD information from the UK Biobank was used (**Supplementary Notes**). All variants were aligned to the same allele as the UK Biobank and any variant with MAF that differed by > 5% were excluded from the fine-mapping analysis. Prior standard deviation for FINEMAP was set to 0.058 (**Supplementary Notes**) and shotgun stochastic search algorithm was used (--sss option) with the maximum number of causal variants in a locus set to five (default). Five annotations were used for PAINTOR: four annotations were chosen based on the relevance of the annotation with the European gout GWAS summary statistics (**Supplementary Notes**) and one annotation was included to give more weight to missense variants. PAINTOR was run with full enumeration (--enum option) assuming single causal variants except for those loci with evidence for multiple signals, in which case a maximum of three causal variants was assumed. For the BF approach, Bayes’ factors of all variants were summed within the fine-mapping region and the posterior probability of each variant being causal in the region was calculated by dividing the variant’s Bayes’ factor with the total Bayes factor of the region. The 99% credible set was determined by including the variant with the highest posterior probability in the region until the cumulative posterior probability of the set was greater than 0.99.

### Identification of compromised loci with SLALOM

To mitigate fine-mapping miscalibration when different genotyping arrays and imputation panels are used in meta-analysis, the summary statistics used for fine-mapping were checked with SLALOM^48^. Conversion-unstable positions (CUPs)^147^ were downloaded and reference LD from gnomAD (build 37) was calculated in the Non-Finnish European population for the European loci and a mixture of African/African American, Latinx/Admixed American, East Asian, and European populations (proportional to the number of samples used in the trans-ancestry meta-analysis) were used for the trans-ancestry loci. A locus was identified as possibly miscalibrated if the locus contained at least one variant with r^2^ > 0.6 with the lead variant (in the relevant reference population) and DENTIST-S^148^ *P*-value was <1×10^−4^ in either the European or the trans-ancestry SLALOM analyses.

### Candidate missense and non-coding variants

A pool of 1466 candidate variants was gathered from lead variants, conditionally associated variants and variants identified from fine-mapping. LD of 1425 non-coding variants was calculated using 1000 Genomes Project data (**Supplementary Notes**), and direct lead or high LD (r^2^ ≥ 0.8) missense and nonsense variants were identified using information from dbSNP (build 155). FATHMM scores for the 1425 variants were queried and the variants were overlayed for activity-by-contact enhancers (**Methods**).

### Co-localization of signals of genetic association with eQTL

Lead SNPs from the European and trans-ancestry analyses, or a proxy if the lead variant was not present in GTEx, were used to query GTEx data for identification of *cis*-eQTL. For all the SNPs with a significant eQTL, a 1Mb region around the lead variant was used for co-localization analysis using the ‘coloc’ R package^64,149^. A posterior probability of >0.5 for H4 (hypothesis 4) was taken as evidence for co-localization, which indicates the scenario that the association between gout and gene expression was due to the same functional variant. We also carried out *trans*-eQTL analysis combining GTEx and publicly available HiC data integrated using the CoDeS3D algorithm^150^ and COLOC^151^ (**Supplementary Notes**).

### Co-localization of signals of genetic association with methylation QTL (meQTL)

The GoDMC data set^39^ was used to first identify all significant DNA methylation QTL (meQTL) within ±1Mb window of the 444 lead European GWAS variants. For each pair of variant-CpG sites, a ±500kb region around the lead GWAS variant was defined and tested for co-localization with the meQTL signal using the ‘coloc’ R package. Regions with < 100 variants with meQTL summary statistics for the CpG sites were excluded and regions with PPH4 ≥ 0.8 were considered as significant.

### Enrichment analysis of transcription factor (TF) binding at meQTL CpG sites

1,544 TF ChIP-seq experiment data sets (344 TFs and 221 cell lines)^40^ were downloaded (**Data Availability**). Eight TF ChIP-seq experiment datasets were excluded to ensure all of the ChIP-seq experiments were from human cell lines (**Supplementary Notes**), leaving 338 unique TFs for enrichment analysis. Positions for 232,477 independent CpG sites used in GoDMC were extracted. For each TF, the total number of CpG sites the TF overlapped (±50bp) within the CpG sites was determined and 520 co-localized meQTL CpG sites identified. 338 two-by-two contingency tables were constructed and Fisher’s exact test was used to determine if there was significant enrichment of TF binding to the meQTL CpG sites.

### Overlap of variants and CpG sites with activity-by-contact (ABC) enhancer region

ABC enhancer-gene connection data for 131 cell types and tissues with ABC-scores ≥ 0.015^35^ was downloaded (**Data Availability**). The positions of candidate non-coding SNPs, lead SNPs that have co-localized eQTL, and CpG sites that have co-localized meQTL were queried in the ABC data to determine whether the SNP and/or the CpG site was within an ABC enhancer region.

### Gene prioritization for gout inflammation

To select the input list of candidate genes, association signal boundaries from all of the loci lists were used to select genes from GENCODE, and co-localized *cis*-/*trans*-eQTL genes not already in the list were added. Satisfying each of the following seven criteria scored the gene one point in the primary prioritization scheme: 1) having an eQTL co-localized with GWAS signal in whole blood; 2) having an eQTL in monocytes (ImmuNexUT^62^ or OneK1K^152^ datasets); 3) within a locus with a co-localized meQTL; 4) within a locus with gout association signal that also genetically associates with one or more of 36 white blood cell traits^78^ (**Table S36** and **Supplementary Notes**); 5) differentially expressed in gout (**Supplementary Notes**); 6) expressed in GTEx whole blood tissue (**Supplementary Notes**); and 7) differentially expressed in monocytes stimulated with MSU crystals and/or lipopolysaccharide (LPS) (LPS vs MSU crystal, LPS vs phosphate-buffered saline control, MSU crystal vs PBS control)^43^. Given that some of the categories used results that were derived using European ancestry and/or trans-ancestry data and could not be applied to all ancestral groups, the prioritisation scores were standardized based on the number of categories to which the gene was applied (depending from which ancestral group the gene was implicated) (**Supplementary Notes**). Following this, within each standardized score category, genes were secondarily ranked according to: 1) one of 47 genes containing a strong candidate missense causal variant (**Table 1**); 2) one of the 385 target genes of the activity-by-contact enhancer; 3) gene with FANTOM5 transcription start site (TSS) closest to the lead SNP. Given that not all genes had TSS information from the FANTOM5^153^ dataset, scores for those genes without TSS information were standardized (Supplementary Notes). Since multiple different SNPs can represent a specific locus (due to different lead SNP across ancestry and/or sex) the highest scoring SNP-gene row was selected to represent the gene.

### Mendelian randomization analysis of CHIP vs gout

GWAS summary statistics for overall CHIP, *DNMT3A* CHIP and *TET2* CHIP were downloaded and filtered to only include variants with *P* < 1×10^−6^, variants in the European meta-analysis for gout, and variants with MAF > 1% in Europeans. For each GWAS, variants were then extracted from the UK Biobank before LD pruning to keep only variants at r^2^ < 0.2 using 50 kb windows, testing every 5^th^ variant (default parameters using PLINK 1.9b6.10). Summary statistics for these variants were extracted for all three CHIP classes along with gout. These summary statistics were used in a two-sample Mendelian randomization analysis, performed using the MendelianRandomization package in R 4.2.1. Beta and standard error values were transformed into log odds ratios and standard errors for each CHIP GWAS. For each CHIP trait, MR was run using three different models: weighted median, inverse variance-weighted and MR-Egger regression.

### Drug repurposing analysis

We applied GREP^63^ to perform the drug repositioning analysis using 476 significant genes in the MAGMA analysis (P < 0.05 / (17,225 genes) = 2.9×10^−6^). GREP uses Fisher’s exact tests to examine whether the 476 genes were enriched in genes targeted by drugs in a clinical indication category: Anatomical Therapeutic Chemical Classification System (ATC) or International Classification of Diseases 10 (ICD10) diagnostic code. ATC has 14 anatomical categories, which are further categorized into 85 detailed classes. GREP lists potentially repositionable drugs targeting the gene set. The threshold for significant enrichment was set as P = 0.05/10, 0.05/85, and 0.05/221 in the ATC large set, the ATC detailed set, and the ICD10 set, respectively, adjusted by the Bonferroni correction for the number of categories. The ATC large set has 14 anatomical categories (e.g. M: MUSCULO-SKELETAL SYSTEM), which are further categorized into ~90 detailed classes (e.g. M04: ANTIGOUT PREPARATIONS) in the ATC detailed set.

### Experimental studies

#### Analysis of single cell RNAseq prostate data

FASTQ files from two single cell RNAseq analyses in prostate tissue involving 3^154^ and 6^155^ samples (3 × 2 zones) were downloaded and analyzed using CellRanger v6.1.0 and Seurat v4.1.1 in R 4.0.2. For each pair of FASTQ files (containing forward and reverse reads respectively), the CellRanger count was used to produce a read count matrix for each sample. The CellRanger count output was read into R using Seurat, then a quality control procedure applied to ensure that cells had less than 20% mitochondrial reads and had greater than 80% complexity (log10 gene count / UMI). Genes with fewer than 10 reads across all cells of each sample were not analyzed. Following quality control, read counts were scaled and transformed using the SCTransform function from Seurat. Samples were then integrated for each study using Seurat, with Henry et al. data^155^ analyzed separately for the peripheral and transition/central zones. Uniform manifold approximation and projection (UMAP) for dimensionality reduction were performed for each sample, then cells were clustered prior to cluster identification. Expression of genes of interest within each cluster was done using the VlnPlot function from Seurat for each study.

#### Measurement of xanthine oxidoreductase (XOR) activity and urate levels in mice

C57Bl/6J mice were housed in the West Virginia University facility in specific pathogen free conditions. All experiments used only 7-week old male mice and were approved by the WVU Institutional Animal Care and Use Committee. Urate concentration was measured in surgically-dissected liver, prostate and seminal vesicle using HPLC coupled to electrochemical detection (ESA Coul-Array System) and expressed per mg protein as previously described^66^. XOR ^112^1 3212321activity (1 Unit = 1 µ mol urate/min) was also assessed using reverse-phase HPLC coupled to electrochemical detection similar to above for urate but with preincubation with xanthine with and without allopurinol to determine contributions directly associated to XOR activity versus existing urate levels ^66^.

## Data availability

The full GWAS summary statistics for the 23andMe discovery data set will be made available through 23andMe to qualified researchers under an agreement with 23andMe that protects the privacy of the 23andMe participants. Datasets will be made available at no cost for academic use (https://research.23andme.com/collaborate/#dataset-access/); 1000 Genomes Project Phase 3 data was downloaded from http://ftp.1000genomes.ebi.ac.uk/vol1/ftp/; information on UK Biobank cohort can be viewed at https://www.ukbiobank.ac.uk/; GWAS Catalog (https://www.ebi.ac.uk/gwas/); GTEx data were downloaded from https://gtexportal.org/home/datasets; variant information from dbSNP was downloaded from https://ftp.ncbi.nih.gov/snp/latest_release/VCF/GCF_000001405.25.gz; CUPs used for SLALOM were downloaded from https://github.com/cathaloruaidh/genomeBuildConversion; GoDMC methylation QTL data were downloaded from http://mqtldb.godmc.org.uk/downloads; RELI transcription factor ChIP-seq data https://tf.cchmc.org/external/RELI/RELI_public_data.tar.bz2; baseline LD score v1.1, cell-type specific, and cell-type group annotations were downloaded from https://alkesgroup.broadinstitute.org/LDSCORE/; functional annotations for PAINTOR were downloaded from https://ucla.box.com/s/x47apvgv51au1rlmuat8m4zdjhcniv2d; LD score regression summary statistics of UK Biobank traits were downloaded from https://nealelab.github.io/UKBB_ldsc/downloads.html; ABC enhancer-gene connection data were downloaded from ftp://ftp.broadinstitute.org/outgoing/lincRNA/ABC/AllPredictions.AvgHiC.ABC0.015.minus150.ForABCPaperV3.txt.gz); FATHMM scores for non-coding variants were obtained from http://fathmm.biocompute.org.uk/; ImmuNexUT eQTL data were downloaded from https://humandbs.biosciencedbc.jp/en/hum0214-v6; OneK1K eQTL data were downloaded from https://onek1k.s3.ap-southeast-2.amazonaws.com/onek1k_eqtl_dataset.zip; Henry *et al.*^155^ prostate single cell RNA-seq FASTQ data were downloaded from https://www.gudmap.org/chaise/record/#2/RNASeq:Study/RID=W-RAHW; Crowley *et al.*^154^ prostate single cell RNA-seq count matrices were downloaded from GEO (GSE150692); CHIP summary statistics were downloaded from https://ftp.ebi.ac.uk/pub/databases/gwas/summary_statistics/GCST90102001-GCST90103000/; Susztak Kidney Biobank (https://susztaklab.com/); a list of differentially expressed genes in stimulated monocytes can be obtained from Table S2 in the original paper^43^; GWAS data for white blood cell traits used in gene prioritization analysis (**Table S30**) (https://www.ebi.ac.uk/gwas/); FANTOM5 TSS data were downloaded from https://fantom.gsc.riken.jp/5/datafiles/phase1.3/extra/TSS_classifier/; HaploReg is available at https://pubs.broadinstitute.org/mammals/haploreg/haploreg.php; GeneHancer tracks were accessed through USCS; pathway analysis websites were https://david.ncifcrf.gov/ and https://maayanlab.cloud/Enrichr/.

## Coding tools availability

Code for “Locus Zoom-like plots” is available at https://github.com/Geeketics/LocusZooms; PLINKv1.9 is available at https://www.cog-genomics.org/plink/; GATK is available at https://gatk.broadinstitute.org/hc/en-us; FINEMAP is available at http://www.christianbenner.com/; PAINTOR is available at https://github.com/gkichaev/PAINTOR_V3.0; LD score regression is available at https://github.com/bulik/ldsc; cov-LD score regression is available at https://github.com/immunogenomics/cov-ldsc; SLALOM is available at https://github.com/mkanai/slalom; CoDeS3D is available at https://github.com/alcamerone/codes3d; GCTA-COJO is available at https://yanglab.westlake.edu.cn/software/gcta/; GREP is available at https://github.com/saorisakaue/GREP.

## Ethics statement

For the 23andMe Inc. Cohort participants provided informed consent and participated in the research online, under a protocol approved by the external AAHRPP-accredited IRB, Ethical and Independent Review Services (www.eandireview.com). The UK Biobank (UKBB) was undertaken with ethical approval from the North West Multi-Centre Research Ethics Committee of the UK. GlobalGout obtained ethical approval from the following committees: the New Zealand Multiregional Ethics Committee (MEC05/10/130); the Northern Y Region Health Research Ethics Committee (Ngāti Porou Hauora Charitable Trust study; NTY07/07/074); Research and Ethics Committee, Repatriation General Hospital, South Australia (32/08); Research Ethics Committee, University of New South Wales; Ethikkommission, Technische Universität Dresden (EK 8012012); South East Scotland Research Ethics Committee (04/S1102/41); Commission Cantonale (VD) D’éthique de la Recherche sur l’être Humain, Université de Lausanne; Commissie Mensgebonden Onderzoek regio Arnhem—Nijmegen; Partners Health Care System Institutional Review Board.hTe Institutional Review Board of the Kaiser Foundation Research Institute provided ethical approval for the Kaiser Permanente sample set. The FinnGen study was approved by theCoordinating Ethical Committee of the Hospital District of Helsinki and Uusimaa. The ethics review board at the Affiliated Hospital of Qingdao University approved the study in China. In Japan ethical approvals were provided by the institutional ethical committees of the National Defense Medical College, Nagoya University and RIKEN. The Korean Association Resource (KARE) was approved by the institutional review board of the Korea National Institute of Health. The FAST study and Generation Scotland was ethically approved by the UK Multi-Centre Research Ethics Committee (Reference number: 2011-001883-23) and the NHS Tayside Committee on Medical Research Ethics (REC Reference Number: 05/Sl401189).

## Data Availability

All data produced in the present study are available upon reasonable request to the authors

## Acknowledgements

We would like to thank the research participants and employees of 23andMe for making this work possible. The following members of the 23andMe Research Team contributed to this study: Stella Aslibekyan, Adam Auton, Elizabeth Babalola, Robert K. Bell, Jessica Bielenberg, Katarzyna Bryc, Emily Bullis, Daniella Coker, Gabriel Cuellar Partida, Devika Dhamija, Sayantan Das, Sarah L. Elson, Nicholas Eriksson, Teresa Filshtein, Alison Fitch, Kipper Fletez-Brant, Pierre Fontanillas, Will Freyman, Julie M. Granka, Karl Heilbron, Alejandro Hernandez, Barry Hicks, David A. Hinds, Ethan M. Jewett, Yunxuan Jiang, Katelyn Kukar, Alan Kwong, Keng-Han Lin, Bianca A. Llamas, Maya Lowe, Jey C. McCreight, Matthew H. McIntyre, Steven J. Micheletti, Meghan E. Moreno, Priyanka Nandakumar, Dominique T. Nguyen, Elizabeth S. Noblin, Jared O’Connell, Aaron A. Petrakovitz, G. David Poznik, Alexandra Reynoso, Morgan Schumacher, Anjali J. Shastri, Janie F. Shelton, Jingchunzi Shi, Suyash Shringarpure, Qiaojuan Jane Su, Susana A. Tat, Christophe Toukam Tchakouté, Vinh Tran, Joyce Y. Tung, Xin Wang, Wei Wang, Catherine H. Weldon, Peter Wilton, Corinna D. Wong.

We thank the participants and staff of the UK Biobank study for their important contributions. This research has been conducted using the UK Biobank resource under application number 12611. We want to acknowledge the participants and investigators of the FinnGen study. This study was supported by grants from the New Zealand Health Research of New Zealand (08/075, 11/1075, 14/527), Arthritis New Zealand, Lottery Health New Zealand, JSPS KAKENHI Grant Numbers (20H00566, 21KK0173, 17H04128, 22H00476, 20K23152, 21H03350, 221S0002, 16H06279 [PAGS], 16H06277 [CoBiA], 20H00568), AMED (JP21gm4010006, JP22km0405211, JP22ek0410075, JP22km0405217, JP22ek0109594), JST Moonshot R&D (JPMJMS2021, JPMJMS2024), Takeda Science Foundation, Bioinformatics Initiative of Osaka University Graduate School of Medicine, the Gout and Uric Acid Foundation of Japan and the Ministry of Defense of Japan, the National Natural Science Foundation of China (82220108015) and National Key Research and Development Program of China (2022YFE0107600; 2022YFC2503300). Generation Scotland received core support from the Chief Scientist Office of the Scottish Government Health Directorates [CZD/16/6] and the Scottish Funding Council [HR03006] and is currently supported by the Wellcome Trust [216767/Z/19/Z]. Genotyping of the GS:SFHS samples was carried out by the Genetics Core Laboratory at the Edinburgh Clinical Research Facility, University of Edinburgh, Scotland and was funded by the Medical Research Council UK, the Wellcome Trust (Wellcome Trust Strategic Award “STratifying Resilience and Depression Longitudinally” (STRADL) Reference 104036/Z/14/Z) and by MRC University Unit grant MC_UU_00007/10 (QTL in Health and Disease programme).

## ^&^Japan Gout Genomics Consortium (Japan Gout)

Ituro Inoue^75^, Toru Shimizu^76^, Hiroshi Ooyama^77^, Keiko Ooyama^77^, Mitsuo Nagase^78^, Yuji Hidaka^79^, Kenji Wakai^80^, Yohko Nakamura^81^, Hiroto Narimatsu^82^, Kiyonori Kuriki^83^, Sadao Suzuki^84^, Hidemi Ito^85^, Asahi Hishida^80^, Takashi Tamura^80^, Yoshikuni Kita^86^, Teruhide Koyama^87^, Kokichi Arisawa^88^, Hiroaki Ikezaki^89^, Keitaro Tanaka^90^, Chihaya Koriyama^91^, Yuzo Takada^92^, Takahiro Nakamura^93^, Miki Ueno^94^, Toshimitsu Ito^95^, Satoko Iwasawa^96^, Satoko Suzuki^96^, Yuka Miyoshi^96^, Hiroshi Nakashima^96^, Yutaka Sakurai^96^, and Masashi Tsunoda^96^.

^75^Human Genetics Laboratory, National Institute of Genetics, Shizuoka, Japan

^76^Midorigaoka Hospital, Osaka, Japan

^77^Ryougoku East Gate Clinic, Tokyo, Japan

^78^Nagase Clinic, Tokyo, Japan

^79^Akasaka Central Clinic, Tokyo, Japan

^80^Department of Preventive Medicine, Nagoya University Graduate School of Medicine, Aichi, Japan

^81^Cancer Prevention Center, Chiba Cancer Center Research Institute, Chiba, Japan

^82^Cancer Prevention and Control Division, Kanagawa Cancer Center Research Institute, Kanagawa, Japan

^83^Laboratory of Public Health, Division of Nutritional Sciences, School of Food and Nutritional Sciences, University of Shizuoka, Shizuoka, Japan

^84^Department of Public Health, Nagoya City University Graduate School Medical Science, Aichi, Japan

^85^Division of Cancer Information and Control, Aichi Cancer Center Research Institute, Aichi, Japan

^86^Faculty of Nursing Science, Tsuruga Nursing University, Fukui, Japan

^87^Department of Epidemiology for Community Health and Medicine, Kyoto Prefectural University of Medicine, Kyoto, Japan

^88^Department of Preventive Medicine, Institute of Health Biosciences, the University of Tokushima Graduate School, Tokushima, Japan

^89^Department of General Internal Medicine, Kyushu University Hospital, Fukuoka, Japan

^90^Department of Preventive Medicine, Faculty of Medicine, Saga University, Saga, Japan

^91^Department of Epidemiology and Preventive Medicine, Graduate School of Medical and Dental Sciences, Kagoshima University, Kagoshima, Japan

^92^Faculty of Medical Science, Teikyo University of Science, Tokyo, Japan

^93^Laboratory for Mathematics, National Defense Medical College, Saitama, Japan

^94^Division of Nursing, National Defense Medical College, Saitama, Japan

^95^Department of Internal Medicine, Self-Defense Forces Central Hospital, Tokyo, Japan

^96^Department of Preventive Medicine and Public Health, National Defense Medical College, Saitama, Japan

